# Genome-resolved quantitative metagenomics of nontuberculous mycobacteria and their associated phages and plasmids in chloraminated drinking water

**DOI:** 10.64898/2026.06.11.26355408

**Authors:** Soojung Lee, Kathryn Langenfeld, Sarah Potgieter, Sadia Marjia Ferdous, Shruti Vasagiri, G. Eric Bastien, Melissa Duhaime, Krista Wigginton, Lutgarde Raskin, Bridget Hegarty

## Abstract

Nontuberculous mycobacteria (NTM) are opportunistic pathogens of growing concern in drinking water, yet the phages and plasmids associated with them remain uncharacterized. We used genome-resolved quantitative metagenomics in three buildings on a chloraminated distribution system. Total bacterial metagenome-assembled genome (MAG) and viral population concentrations averaged 8.41 × 10^7^ and 8.00 × 10^8^ copies/L, respectively, while the total concentration of the seven NTM MAGs averaged 4.01 × 10^5^ copies/L. Converging genomic evidence supported the identification of four viral MAGs (vMAGs) as putative mycobacteriophages. Three of these vMAGs encoded lysin B, a mycolylarabinogalactan esterase that cleaves the linkage between mycolic acids and arabinogalactan in the cell envelope, and three encoded a WhiB-family transcriptional regulator. NTM concentrations were highest at the site with the lowest bacterial and viral diversity. Combined four-vMAG concentrations were positively associated with total NTM concentrations within sites, although the association across all nine samples was not significant. NTM plasmids carried ESX secretion and metal efflux genes at higher proportions than NTM chromosomes. NTM carried more steroid degradation genes than co-occurring Actinomycetota. These findings reveal site-specific NTM–phage concentration relationships and identify phage- and plasmid-associated functions with potential implications for NTM–phage interactions and environmental adaptation.

**Table of Contents (TOC):** 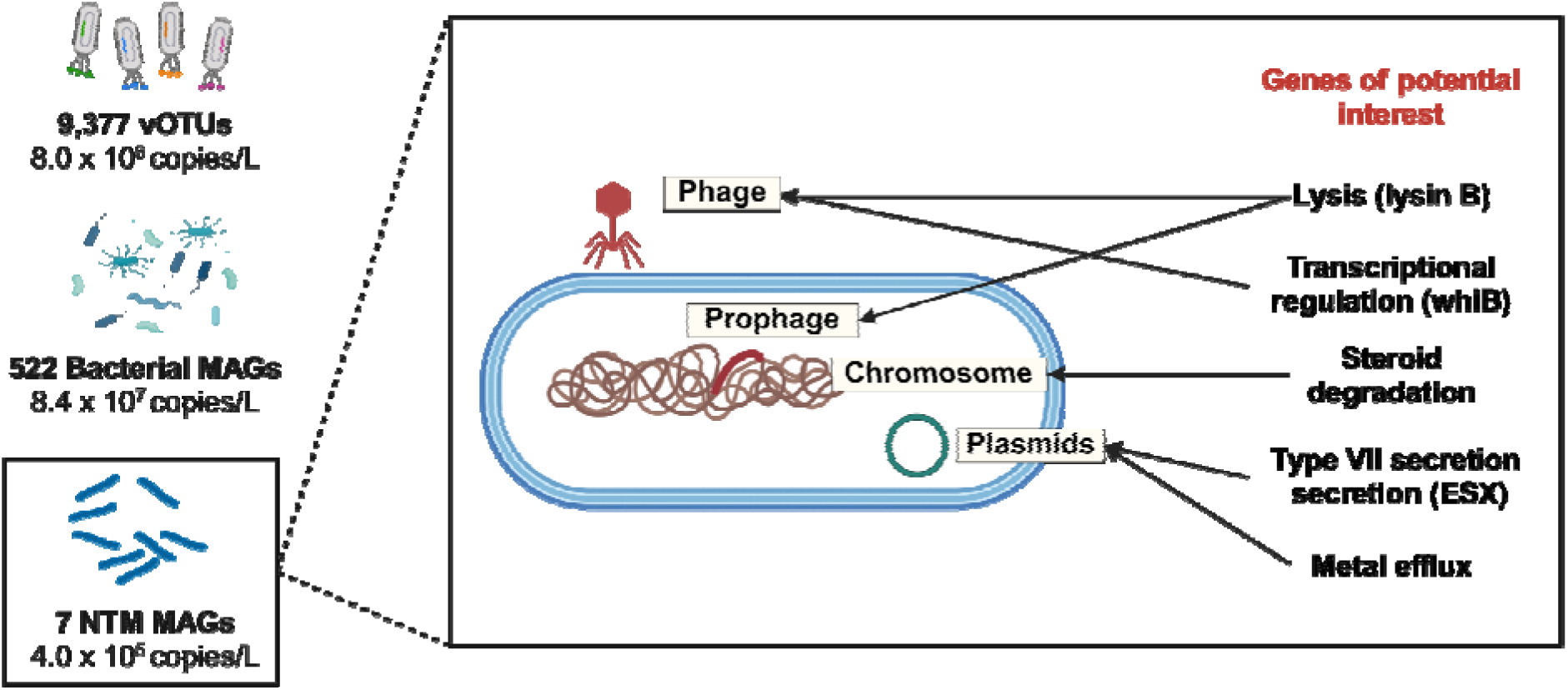

## Introduction

Drinking water can harbor opportunistic pathogens that pose risks to human health and contribute to substantial economic burdens. In the United States, drinking water exposures account for approximately 40% of hospitalizations and 50% of reported deaths associated with waterborne disease, with annual healthcare costs estimated at $1.39 billion^1^. Nontuberculous mycobacteria (NTM) alone account for approximately 46% of this total economic burden^1^. Specific NTM species, including *Mycobacterium abscessus* and *Mycobacterium avium*, can cause pulmonary and soft tissue infections^2^. These species were included in the U.S. Environmental Protection Agency’s Contaminant Candidate List 5 (CCL 5)^3^, which prioritizes unregulated drinking water contaminants for monitoring, research, and possible future regulation. Draft CCL 6 further added the “pathogenic waterborne mycobacteria group”, including nine NTM species^4^. NTM persistence is supported by a robust, mycolic acid-rich cell envelope^5^, the capacity to form biofilms^2^, and associations with free-living amoebae that provide protection from environmental stress^6^. NTM are detected more frequently and at higher concentrations in chloraminated drinking water distribution systems than in those using free chlorine^2,7^. This proliferation is further amplified within building plumbing, where decay of disinfectant residuals, high surface-to-volume ratios, prolonged stagnation, and higher water temperatures are conducive to microbial growth and biofilm formation. Because building plumbing represents the final critical point of human exposure, quantifying NTM and characterizing the genetic traits they carry at this point is essential for managing NTM in drinking water.

Beyond the intrinsic cellular traits that enable survival and growth in building plumbing, NTM persistence may also be influenced by phages and plasmids. Phage–host interactions range from lytic infection to lysogeny and can have divergent consequences for bacterial populations^8^. Lytic infection can reduce bacterial abundance through cell lysis, and some mycobacteriophages (viruses that infect *Mycobacterium*) encode lysis-associated enzymes, including LysB, which facilitates disruption of the mycobacterial cell envelope^9^. Phage infection can also induce metabolic reprogramming of infected cells (virocells), altering host nutrient acquisition, energy metabolism, and stress responses^10,11^. In drinking water, phages have been reported to carry genes associated with oxidative and nitrogen stress responses^12^. Lysogeny may also support bacterial persistence through prophage-encoded accessory genes^13,14^ and protection against infection by related phages^15^. Plasmids represent another source of accessory genes, including those associated with antimicrobial resistance, virulence, and Type VII secretion (ESX) systems^16,17^. Although putative mycobacteriophages have been detected in chlorinated drinking water^9^, research on these viruses has focused primarily on clinical diagnostics and therapy, including treatment of drug-resistant *M. abscessus* infections^18–21^. However, the prevalence, genetic diversity, and potential ecological roles of NTM-associated phages and plasmids in drinking water systems remain poorly understood.

Existing culture-based methods, amplicon sequencing, and targeted quantitative PCR (qPCR) generally provide either taxon-specific measurements or community-level relative abundances, but rarely link genome-resolved identity to absolute concentration. Because relative abundances depend on community composition, they may not track the absolute concentration of a population, and two populations can appear to change together even when their absolute concentrations do not^22^. A previous study of a *Mycobacterium*-dominated, chlorinated drinking water distribution system characterized viral community composition based on relative abundance^9^. Recently developed qPCR assays targeting major capsid protein genes of selected Caudoviricetes phages infecting *Mycobacterium* spp. demonstrate increasing interest in their environmental quantification, but targeted assays cannot capture the broader diversity of uncharacterized phages or resolve their genomic associations with environmental hosts^23^. NTM, viral populations, and the broader bacterial community have not been quantified simultaneously in absolute terms in the same drinking water samples. Quantitative metagenomics^24^ using spiked-in standards can address this gap by enabling estimates of their absolute concentrations.

Here, we combined genome-resolved metagenomics with absolute quantification to investigate NTM, viral populations, and plasmids in chloraminated building plumbing water. First, we characterized viral populations in drinking water predicted to interact with NTM and tested our hypothesis that mycobacteriophage concentrations would be positively correlated with host concentrations, as greater host abundance may support increased phage replication^25^. Second, we examined how the functional gene content of NTM differs from that of other Actinomycetota in the same samples. Third, we investigated whether NTM-associated plasmids carry genes that expand the functional potential of NTM chromosomes. By integrating absolute concentration measurements, predicted phage–host associations, and functional gene annotations, this study characterizes NTM-associated viral and plasmid gene pools and identifies potential ecological interactions and functional potentials that warrant further investigation.

## Methods

### Source water and water treatment plant

The Ann Arbor water treatment plant has been described previously by Dowdell et al. (2024) and Potgieter et al. (2025)^26,27^. In summary, the plant supplies drinking water to approximately 125,000 residents and produces an average of 15 million gallons per day (5.7 × 10^4^ m^3^/day). The source water consists of a seasonal blend of surface water (80–85%) and groundwater (15–20%). The treatment process involves two-stage lime softening, followed by coagulation, flocculation, and sedimentation. Ozone is applied as the primary disinfectant (approximately 2 mg·min/L) prior to biologically active dual-media filtration using granular activated carbon and sand. Monochloramine (about 3 mg/L as Cl_2_) is applied as the secondary disinfectant before the treated water is distributed.

### Sampling location and sample collection

This study was conducted at three building locations in Ann Arbor, Michigan, U.S. (Sites A, B, and E). These sites are used by the City of Ann Arbor for water quality monitoring and have been included in previous studies^26,27^. Site A was selected because it is located close to the water treatment plant. Sites B and E were selected as they were previously found to have relatively high NTM diversity and concentrations^26^. Site A is a municipal building, whereas Sites B and E are private buildings. Samples were collected from a janitorial closet tap at Site A, a bathroom sink at Site B, and a handwashing sink at Site E (Table S1).

Samples were collected on June 7 (Day 1), June 9 (Day 3), and June 13 (Day 7) in 2022 at approximately 9:00 AM on each sampling day, following the same site order (Site A, then Site B, then Site E). The first 2 L of tap water were collected in sterile polypropylene bottles to represent water from the fixture and the building plumbing (first-draw samples). First-draw sampling was selected to characterize water at the point of use before flushing and to obtain microbial biomass for genome-resolved quantitative metagenomics. All three buildings are non-residential (Table S1) and generally unoccupied overnight and on weekends. Because June 13 was a Monday, the Day 7 samples likely followed weekend stagnation, whereas the Day 1 and Day 3 samples reflected overnight stagnation under routine building operation. However, the exact duration of stagnation and the preceding water-use history at each fixture were not recorded.

From each 2 L sample, 500 mL was concentrated using an automated Concentrating Pipette Select (InnovaPrep, Drexel, MO, USA) with 100 kDa ultrafilters, and the concentrate was eluted with 500 µL phosphate-buffered saline (PBS). The recovery of these filters for bacteriophage T4 was 81 ± 9.7%, measured by droplet digital PCR (Bio-Rad) following a previously described approach^28^. After filtration and elution, the nine first-draw samples were stored at −80 °C.

Physicochemical parameters were measured immediately after sampling, as described by Potgieter et al. (2025)^27^ (Table S2). Temperature, pH, electroconductivity, ORP, and total dissolved solids were measured using a Hanna probe (HI98129 and HI98121; Hanna Instruments, Woonsocket, RI, USA). Total chlorine, monochloramine, free ammonia, total ammonia, orthophosphate, nitrite, and dissolved iron concentrations were quantified using the Hach parallel analyzer with corresponding Chemkeys®. Samples for total organic carbon (TOC) were collected in carbon-free glass vials, acidified with 85% phosphoric acid, and analyzed as non-purgeable organic carbon using a Shimadzu TOC analyzer (TOC-V, Japan).

### DNA extraction

DNA was extracted using a modified protocol of the DNeasy PowerWater Kit (QIAGEN). The modifications included enzymatic treatments with Lysozyme and Proteinase K, the use of chloroform:isoamyl alcohol (24:1), and altered bead-beating steps^29^ (dx.doi.org/10.17504/protocols.io.66khhcw). The extracted DNA was quantified using the Qubit dsDNA High Sensitivity Assay Kit (Thermo Fisher Scientific) in conjunction with the Qubit 4 Fluorometer (Thermo Fisher Scientific). DNA extracts (100 µL) were stored at −80 °C until sequencing (Table S1).

### Short-read sequencing and preparation of metagenome-assembled genomes (MAGs)

Short-read sequencing was performed at the University of Michigan Advanced Genomics Core. Initial quality control was performed with Qubit to measure DNA concentration, NanoDrop for purity assessments, and Agilent Tapestation for fragment size estimation. Library prep was done using the xGen™ ssDNA & Low-Input DNA Library Prep Kit (Integrated DNA Technologies, Inc., Coralville, Iowa). One sample (SiteA_UF_fd_7) was sequenced on a NextSeq P2 600-cycle flow cell due to low biomass. The remaining samples were sequenced on a NovaSeq 6000 SP 500-cycle flow cell. Both resulted in 250 bp reads. Sequencing data were processed using the workflow established by Vosloo et al. (2021)^30^, including sequence processing, de novo co-assembly, and MAG reconstruction, as detailed in Text S1.

### Long-read sequencing and processing

Long-read sequencing of the first-draw samples was unsuccessful because of insufficient DNA yield. Instead, three full-flush samples from Site A, collected on Days 1, 7, and 8, were used for long-read sequencing (Text S2). For first-draw samples, 500 mL of each of the 2 L water samples was processed for microbial concentration, yielding DNA concentrations of 0.21–2.06 ng/µL (Table S1). In contrast, the three full-flush samples, collected on 0.22 µm Sterivex filters, yielded DNA concentrations of 29.6, 25.9, and 32.7 ng/µL.

The long-read data were used to provide sequence-level support for regions of the short-read viral assemblies. Only long-read assemblies also detected in the short-read data were retained for downstream analysis. Long-read sequencing, sequence processing, and assembly are detailed in Text S3.

### Quantitative metagenomic analysis

Before library preparation, synthetic dsDNA and ssDNA standards were spiked into each sample at known abundances to relate observed relative abundances to absolute abundances^24,31^. For all samples except SiteB_UF_fd_7, Sequins dsDNA metagenomics mix B^31^ and a mix of five ssDNA standards^24^ were added at 1.0% and 0.05% of total DNA mass, respectively (Table S3). In SiteB_UF_fd_7, only 0.2% of Sequins dsDNA metagenomics mix B was spiked into the sample.

The absolute abundances of MAGs and vOTUs were determined using QuantMeta, with previously established thresholds for detection and read-depth variability^24^. Individual MAG absolute abundances were calculated as the mean absolute abundance of each contig normalized by length, with zeroes for contigs below detection. Values were then converted to copies per liter (copies/L) based on the DNA extraction elution volume (100 µL) and the filtered sample volume (Table S1). vOTUs are composed of individual contigs, so the concentrations directly calculated by QuantMeta were accepted as the vOTU concentrations (copies/L).

### Viral sequence identification and host prediction

Viral contigs were identified and clustered into species-level vOTUs, followed by taxonomic and lifestyle characterization. Virus–host associations were predicted using VHIP and iPHoP. Detailed procedures are provided in Text S4.

### Plasmid and prophage identification

Plasmid and prophage sequences associated with NTM MAGs were identified and characterized using sequence-based approaches. Detailed procedures are provided in Text S5.

### Metabolic potential analysis

Functional potential of MAGs, associated plasmids and prophages, and vOTUs was evaluated using KEGG-based annotation and complementary analyses of virulence, antimicrobial resistance, and horizontal gene transfer. Gene and KO abundances were estimated using metagenomic read coverage and QuantMeta-derived MAG abundances. Detailed procedures are provided in Text S6.

### Data analysis

All statistical analyses and visualizations were conducted in RStudio (2023.09.1+494) with R (v4.4.0) unless otherwise noted. Figures were generated using ggplot2 (v3.5.2) unless otherwise noted. Alpha diversity was calculated using Shannon and Simpson indices, and community composition was analyzed by PERMANOVA, distance-based redundancy analysis (dbRDA), and variance partitioning, all using vegan (v2.6-10). Relationships between concentrations were evaluated using Pearson and Kendall rank correlations and, to account for the nested sampling design, linear mixed-effects models fitted with site as a random intercept. Differences among sites were tested using the Kruskal–Wallis test, and gene content was compared between groups of MAGs and between genomic compartments using Wilcoxon and Fisher’s exact tests. All tests were two-sided, statistical significance was defined as *p* < 0.05, and Benjamini–Hochberg corrected values are reported as q. The variables entered into each test, the transformations applied, the software versions, and the families over which corrections were applied are detailed in Text S7.

## Results and Discussion

### Quantitative metagenomics enabled absolute quantification of bacterial and viral populations in drinking water

First-draw samples capture water that has stagnated in building plumbing and provide an exposure-relevant representation of microorganisms present at the point of use. We collected first-draw samples from three buildings^26^ (Sites A, B, and E) within a chloraminated drinking water distribution system on Days 1, 3, and 7 (Table S1). Sampling within a single distribution system over one week enabled examination of short-term variation in microbial community composition and abundance while limiting differences associated with seasonal change. Short-read sequencing recovered 522 medium-quality bacterial MAGs^32^ (completeness ≥50% and redundancy <10%), as well as 9,377 viral operational taxonomic units (vOTUs; >3 kb), representing metagenomic proxies for distinct viral populations clustered by 95% average nucleotide identity (ANI). Of these vOTUs, 414 with a completeness ≥75% were designated as vMAGs (Fig. S2).

We converted the relative abundances of bacterial MAGs and vOTUs to absolute concentrations (copies/L) using a quantitative metagenomics framework^24^ with Sequins dsDNA standards added at 1.0% of library input DNA mass in eight samples and 0.2% in one sample (Table S3). Across the nine samples, absolute concentrations varied more widely than relative abundance (RPKM) for both bacterial MAGs (10.3-fold vs. 1.3-fold) and vOTUs (28.4-fold vs. 9.5-fold; Fig. S3), demonstrating the value of quantitative metagenomics in revealing variation in microbial concentrations that cannot be quantified from relative abundance data alone. The median cumulative concentration of individual viral populations across the nine samples was higher for vMAGs than for all vOTUs (3.54 × 10^5^ vs. 1.10 × 10^5^ copies/L; Fig. 1A), consistent with the greater assembly completeness generally achieved for more abundant viral populations. Total bacterial MAG concentrations averaged 8.41 × 10^7^ ± 5.51 × 10^7^ copies/L, whereas total vOTU concentrations averaged 8.00 × 10^8^ ± 5.39 × 10^8^ copies/L. Bacterial MAG concentrations were approximately 1.6 orders of magnitude higher than the previously reported total bacterial 16S rRNA gene copies measured by qPCR in the same system^33^ (∼2 × 10^6^ copies/L). These differences reflect methodological variation, including differences in DNA extraction methods and metagenomics-based vs. qPCR-based quantification approaches, as well as differences in sampling time and location.

**Figure 1.**
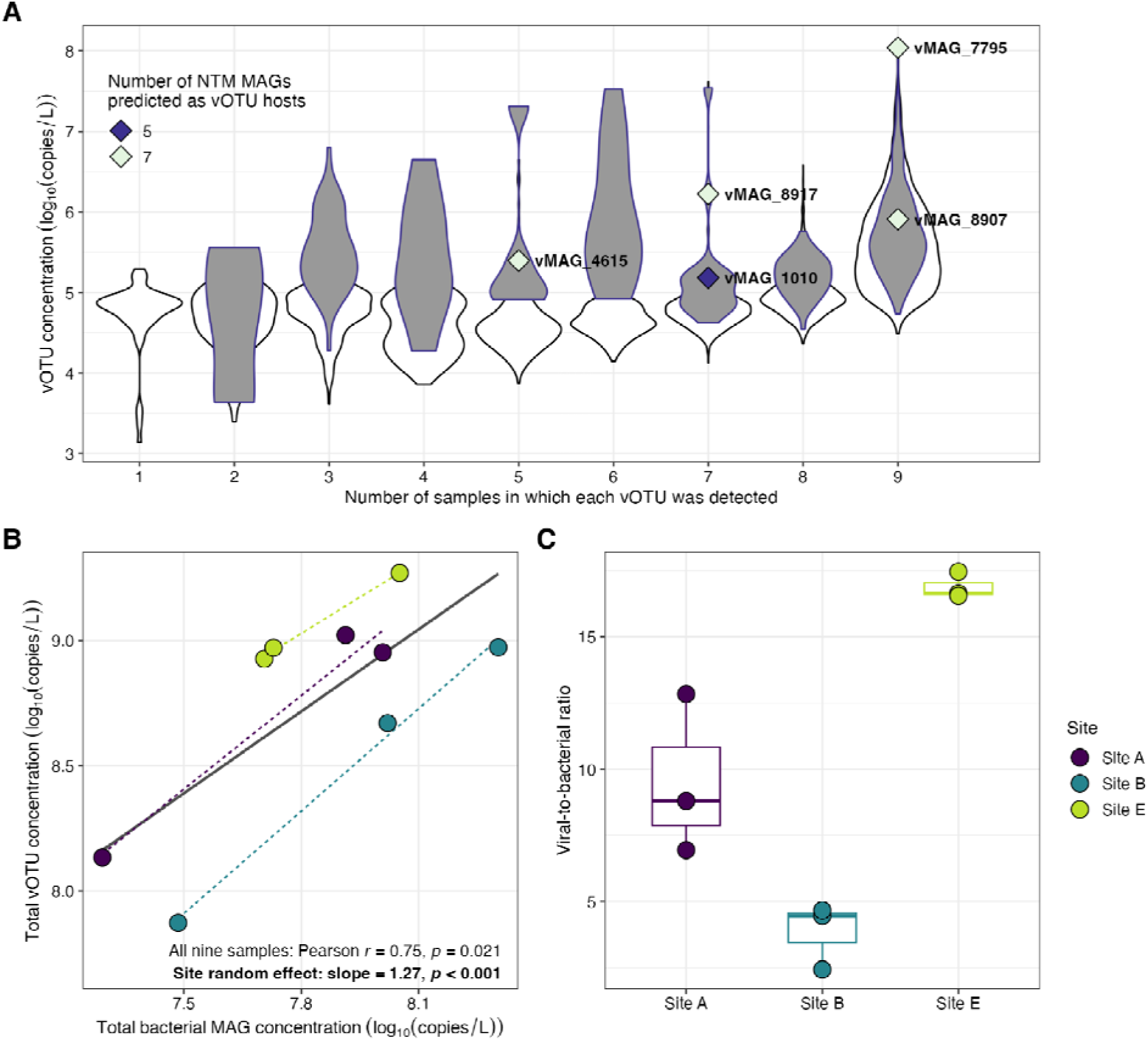
Bacterial MAG and vOTU abundance patterns. (A) Distribution of cumulative absolute concentrations of the 9,377 vOTUs across the number of samples in which they were detected. The white violin plot represents all vOTUs, and the grey violin plot represents the subset of 414 vMAGs with completeness ≥75%. Five vMAGs are highlighted according to the number of NTM MAGs they were predicted to interact with based on VHIP (Table S4). (B) Total vOTU and bacterial MAG concentrations in each sample, colored by sample site. The solid line represents the linear regression across all nine samples (Pearson r = 0.75, *p* = 0.021). Dashed lines represent separate regressions within each site. Because the nine samples are nested within three buildings, the relationship was also fitted with site as a random effect (linear mixed-effects, slope = 1.27, *p* < 0.001). (C) Viral-to-bacterial ratios, calculated as total vOTU concentration divided by total bacterial MAG concentration, across the nine samples. Points are colored by sample sites.

Bacterial MAG and vOTU concentrations were positively associated across the nine samples (Pearson r = 0.75, *p* = 0.021; Fig. 1B). Positive associations were also observed within each building, and the relationship remained significant when sites were included as a random effect (linear mixed-effects, *p* < 0.001). However, the small number of observations per site (n = 3) limits the interpretation of these relationships. The viral-to-bacterial ratio, calculated as the total vOTU concentration divided by the total bacterial MAG concentration, averaged 10.1 ± 5.9 (range: 2.4–17.5; Fig. 1C). To our knowledge, this is one of the first metagenomics-based estimates of an absolute viral-to-bacterial ratio in drinking water. The observed ratios show convergence with the range reported for some marine environments based on particle-based viral-to-bacterial ratios^34^, although direct comparisons are constrained by differences in viral recovery and quantification approaches (particle-based vs. our genome equivalents). Previous drinking-water metagenomic work found that viral contigs represented 9% of contigs longer than 3 kb but did not quantify viral abundance relative to bacterial abundance^12^.

We compared the short-read viral assemblies with long-read assemblies generated from full-flush samples from Site A. The most abundant short-read vMAG, vMAG_7795 (65.4 kb; Tables S4 and S5; 1.22 × 10^7^ ± 8.31 × 10^6^ copies/L), was also represented in the long-read dataset, with a 22.2 kb long-read contig mapped to vMAG_7795, supporting the continuity of the aligned region. Overall, approximately one-third of the short-read viral populations were detected in the long-read assemblies (Fig. S2). Long-read sequencing generally requires higher DNA quality^35^, whereas the extraction protocol used here was optimized to enhance lysis of hard-to-lyse cells in low-biomass drinking water samples, resulting in substantial DNA shearing and no long reads >25 kb. The limited overlap may reflect both DNA shearing and differences in viral community composition between first-draw (short-read) and full-flush (long-read) samples, although their relative contributions cannot be determined from these data. For this reason, we have focused only on long-read contigs found in the short-read data, as this provides independent support for the integrity of selected assembled regions.

Most vOTUs (98.6%) were assigned to the class Caudoviricetes, consistent with a recent drinking water virome study, in which 96% of vOTUs were assigned to the same class^9^. The second most abundant class was Megaviricetes (0.89%). Most vOTUs were unclassified at the family level (Fig. S4), consistent with studies of aquatic and drinking-water viromes^9,12,36–38^ and indicating that drinking-water viral diversity remains poorly represented in current reference databases. Among classified vOTUs, Zobellviridae and Inoviridae were abundant (Fig. S4). Detection of Inoviridae, a family of single-stranded DNA viruses, demonstrates that our library preparation approach recovered both ssDNA and dsDNA viruses: to our knowledge, the first report of ssDNA viruses in drinking water.

Among the 522 bacterial MAGs, seven were assigned to NTM by GTDB-Tk (Table S6). Of these, five were included in a maximum-likelihood phylogenetic tree constructed from single-copy marker genes using GToTree (Fig. S5), as the remaining two lacked sufficient marker genes. We categorized these seven NTM MAGs according to clinical risk group definitions^39^.

Three were categorized as Risk Group 2 (opportunistic pathogens capable of causing disease but mostly treatable): *M. avium*, *Mycobacterium phocaicum*, and *Mycobacterium arupense*. The remaining four, *Mycobacterium gordonae*, *Mycobacterium paragordonae*, *Mycobacterium gadium,* and *Mycobacterium llatzerense*, were classified as Risk Group 1 (species rarely associated with human or animal disease). Among the Risk Group 2 taxa, *M. avium* exhibited 98.86% ANI relative to the reference genome of *M. avium* subsp. avium (GCF_009741445.1), though subspecies-level assignment from ANI alone should be interpreted with caution because clinical relevance depends on additional genomic and phenotypic contexts. The combined concentration of the seven NTM MAGs averaged 4.01 × 10^5^ ± 3.52 × 10^5^ copies/L, consistent with a prior qPCR study at the same sites, which reported 10^4^ to 10^6^ gene copies/L^26^. Quantitative metagenomics expanded NTM detection relative to prior culture- and PCR-based approaches, recovering greater taxonomic diversity (Fig. S6) and enabling species-level concentration comparisons across sites.

### Host genomic GC content contributes to broad VHIP-predicted NTM–phage associations

Virus–Host Interactions Predictor^40^ (VHIP) was applied to predict virus–host interaction networks. VHIP is a machine-learning tool with a reported MAG-level accuracy of 87.8% for classifying virus–host pairs as infection or non-infection. It uses bacterial, archaeal, and viral genome sequences to compute coevolutionary and sequence-based features, including k-mer distances, GC content differences, and sequence homology. VHIP predicted extensive host associations across the microbial community. All 414 vMAGs were predicted to interact with at least one bacterial MAG in our dataset (Fig. S7); 92% of vMAGs were predicted to interact with at least one of the seven NTM MAGs, and 28% were predicted to interact with all seven (Fig. 2A). vMAGs predicted to interact with more NTM MAGs also tended to be linked to more bacterial hosts overall (Pearson, r = 0.54, *p* < 0.001; Fig. 2B). These patterns suggest that predicted NTM-associated vMAGs are not necessarily NTM-specific, as many were also linked to diverse non-NTM bacterial hosts. Cross-phylum infection has been experimentally demonstrated for several phages and has also been inferred using proximity-ligation approaches in environmental communities^41–44^. As VHIP’s predictions are based on genomic features that may persist longer than infectivity, our predictions may indicate that these vMAGs have evolved to rapidly change hosts rather than indicate current broad infectivity, an ability that could be advantageous in low-biomass, oligotrophic building-plumbing environments. However, experimental or proximity-based validation will be needed to establish cross-phylum infection.

**Figure 2.**
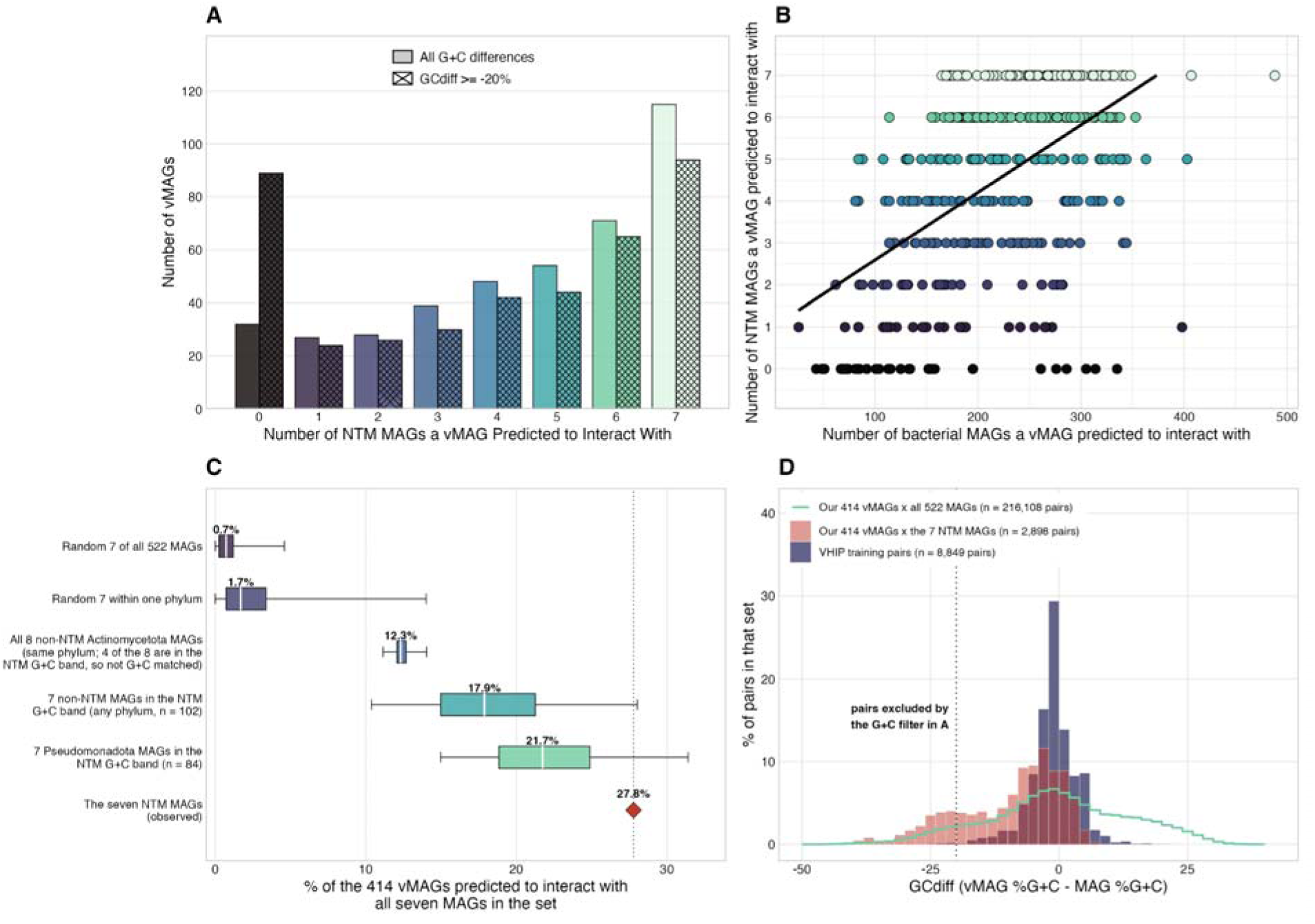
Predicted NTM–phage interaction patterns and the contribution of host GC content. (A) Number of vMAGs predicted by VHIP to interact with each of the seven NTM MAGs, including vMAGs with no predicted NTM host (zero). For each count, the left bar uses all virus– host pairs and the right bar excludes pairs whose GC difference falls below −20% (panel D). (B) Number of predicted NTM MAG hosts against the total number of predicted bacterial MAG hosts, one point per vMAG, using all pairs (Pearson r = 0.54, *p* < 0.001). (C) Percentage of the 414 vMAGs predicted to interact with all seven MAGs of a set, for five comparison sets and for the observed seven NTM MAGs (red diamond). Boxes span the interquartile range and whiskers the 2.5th and 97.5th percentiles across 2,000 random sets; the white line and the number above each box are the median. The Actinomycetota row is not a random draw but all eight possible sets of seven from the eight non-NTM Actinomycetota MAGs, and four of those eight fall inside the NTM GC range, so that row is a same-phylum rather than a GC-matched comparison. (D) Distribution of the GC difference between virus and host, for VHIP’s training pairs, for our vMAG–NTM pairs and for all our vMAG–bacterial MAG pairs. The dotted line marks the −20% cut applied to the right bar of each pair in panel A.

We examined whether this extensive predicted host sharing was associated with the narrow, high-GC-content range occupied by the seven NTM MAGs (65.0–69.2%; Fig. S8A, B). Across 2,000 random sets of seven bacterial MAGs drawn from the 522 MAGs, a median of 0.7% of vMAGs were predicted to interact with all seven (Fig. 2C). This proportion increased to 17.9% for sets drawn from the 102 non-NTM MAGs within the NTM GC range and to 21.7% for sets drawn from the 84 Pseudomonadota MAGs within that range. Although these results suggest that GC content contributes to extensive predicted host sharing, the higher proportion observed for the seven NTM MAGs (28%) suggests that additional genomic features may also contribute. In our study, 19% of the vMAG–NTM pairs had GC differences (GC of vMAG minus GC of bacteria) below -20%, a range poorly represented in VHIP’s training data (Fig. 2D). Excluding these pairs increased the proportion of vMAGs with no predicted NTM host from 8% to 22% (Fig. 2A). GC content is associated with codon usage patterns, which can reflect evolutionary adaptation between phages and their bacterial hosts^45^. Genomic composition may contribute to the extensive predicted phage–host associations observed here, although whether these predictions reflect true broad host ranges requires experimental validation.

### Four vMAGs carry genomic evidence of NTM association

Given the extensive host sharing predicted by VHIP, we examined additional genomic evidence to identify vMAGs with stronger support for NTM association. We used an additional virus–host prediction tool, iPHoP, a machine-learning-based tool that integrates multiple prediction approaches and provides high-confidence genus-level host predictions from viral genome sequences, with an estimated false discovery rate below 10%. Using its default database (Text S4), iPHoP produced 137 genus-level predictions for 121 of the 414 vMAGs, including four assigned to *Mycobacterium* (confidence scores 91.4–99.4; Fig. S9). With the custom database, five high-confidence predictions (scores >90) linked vMAG_4615 to the *M. arupense* MAG and vMAG_8907 to four NTM MAGs in our study (Fig. S9). Fifteen vOTUs shared at least 1 kb of genomic sequence with known mycobacteriophages in the Actinobacteriophage database^46^ (71– 80% nucleotide identity; Table S7). Based on these results, we highlight four vMAGs (vMAG_1010, vMAG_4615, vMAG_8907, and vMAG_8917) as putative mycobacteriophages (Table S4). All four were detected in more than half of the samples (Table S5) and had cumulative concentrations exceeding 10^5^ copies/L (Fig. 1A). In addition, all four retained their predicted NTM host interactions after excluding VHIP-predicted virus–host pairs with GC differences below −20%, as their differences from the seven NTM MAGs ranged from −7.1% to +2.9%.

Among the four candidates, vMAG_1010 and vMAG_4615 had the strongest genomic evidence of NTM association. vMAG_1010 shared sequence similarity with the mycobacteriophages JustASigh and Weirdo19, encoded an Arg tRNA identical to that of the *M. llatzerense* MAG, and was assigned to *Mycobacterium* by iPHoP using the default database (Table S4). vMAG_4615 corresponded to a 54-kb prophage region within the *M. arupense* MAG and shared 99.8% nucleotide identity across the aligned region, and iPHoP predicted its host to be *M. arupense* using both databases (Table S4). vMAG_4615 was the only nearly complete prophage found in an NTM (54.2 kb, 76 genes, seven viral hallmark genes, 97% completeness); four other prophage regions were detected, but their sizes were small, consistent with partial or decaying prophages (Fig. S10). vMAG_8907 and vMAG_8917 also had NTM hosts predicted by iPHoP (vMAG_8907: *M. phocaicum*, *M. arupense*, *M. llatzerense*, and *M. gadium* MAGs, and vMAG_8917: *M. paragordonae* MAG). They also had sequence similarity to known mycobacteriophages (vMAG_8917 to mycobacteriophage Soul22 and vMAG_8907 to mycobacteriophage Ramsey) (Table S4). Notably, vMAG_8907 encoded a tRNA matching the non-NTM MAG_218 (*Sphingopyxis* sp030145715), which VHIP also predicted as a host, further suggesting the potential broad host range of putative mycobacteriophages.

These four vMAGs carried 77 ± 10 predicted genes each, of which 39 ± 5% received functional annotations (Table S8). All exhibited a canonical siphovirus-like genome organization^47^ (Fig. 3). In addition, 17 ± 5% of their VOG-annotated genes had best hits from mycobacteriophage L5 or D29 (Table S8), compared with 4 ± 6% among the other vMAGs (Wilcoxon rank-sum test, *p* = 0.001).

**Figure 3.**
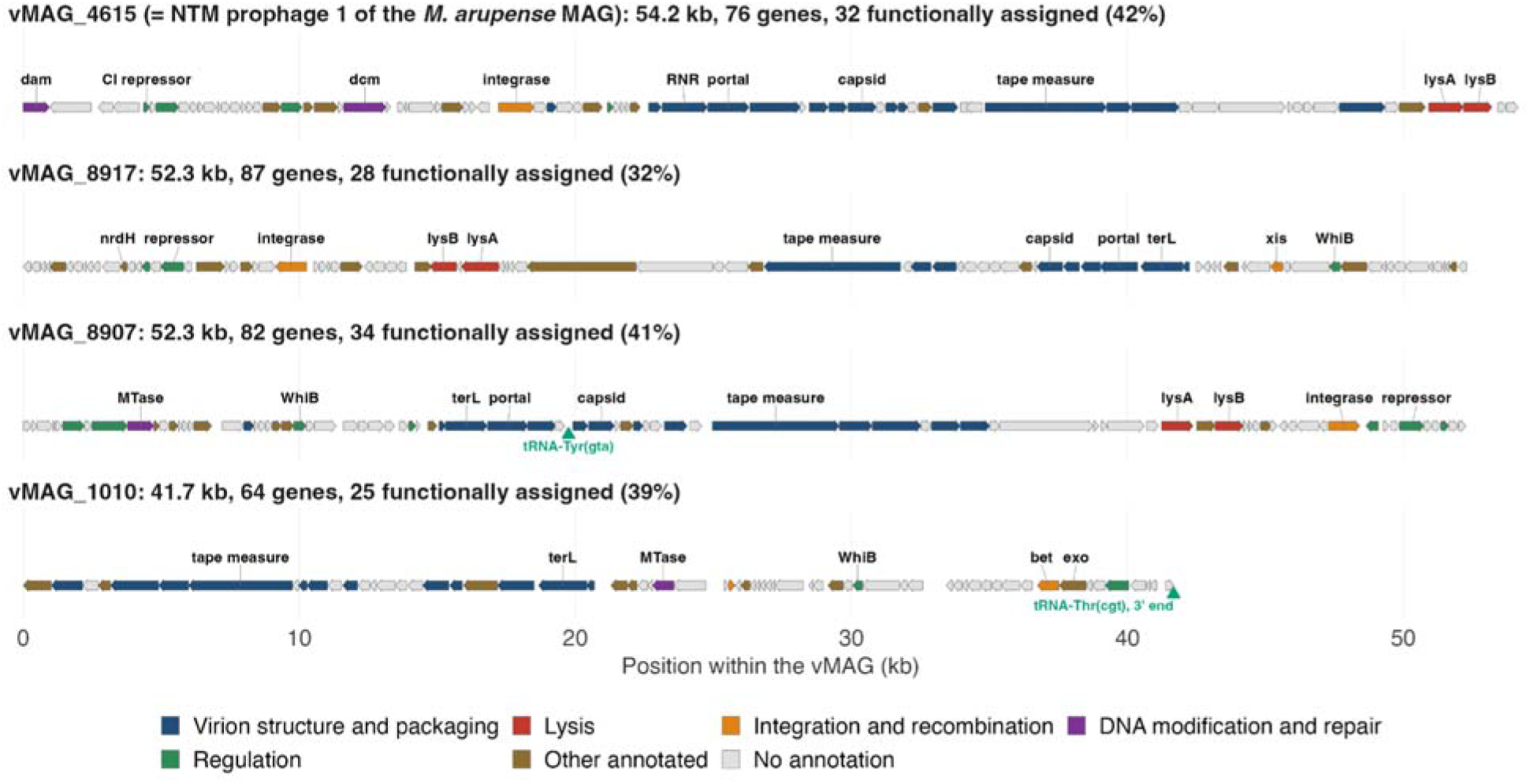
Gene maps of the vMAGs recovered from the NTM MAGs. Each arrow is a predicted gene and is colored by the functional category. Module assignments were made from combined annotations of geNomad, VIBRANT (Pfam and VOG), and GhostKOALA (KEGG Orthology), and genes matching only hypothetical proteins are shown as unannotated. tRNA are labeled in green. Strip headings give the host MAG, the length of the region, and the number of predicted genes.

Two genes identified in these four vMAGs, but not in the other vMAGs, were lysin B and a WhiB-family transcriptional regulator (Fig. 3). Three vMAGs encoded lysin B (vMAG_4615, vMAG_8907, and vMAG_8917), a mycolylarabinogalactan esterase that cleaves the linkage between mycolic acids and arabinogalactan in the mycobacterial cell envelope^48^. Its presence is consistent with the potential to infect mycolic acid-containing bacteria. vMAG_4615 encoded adjacent lysin A and lysin B genes, whereas vMAG_8907 and vMAG_8917 also carried peptidoglycan-hydrolyzing enzymes or lysin A near lysin B (Fig. 3). LysB was also recently reported in three viral populations from a chlorinated drinking water distribution system, including two classified as mycobacteriophages^9^. Furthermore, LysB from mycobacteriophage D29 has demonstrated activity against drug-resistant *Mycobacterium tuberculosis*^49^. Three vMAGs (vMAG_1010, vMAG_8907, and vMAG_8917) also encoded WhiB-family regulators implicated in host cell division and superinfection exclusion in mycobacteriophages^50^ (Fig. 3). A WhiB homolog encoded by mycobacteriophage TM4 has been shown to interfere with host cell division and promote superinfection exclusion^50^. The presence of lysin B and WhiB-family regulators suggests potential applications in phage-based biocontrol of NTM through cell envelope degradation and interference with host cell division, although their specific functions and efficacy remain to be experimentally validated.

Three vMAGs (vMAG_4615, vMAG_8907, and vMAG_8917) encoded an integrase together with a putative repressor or antirepressor (Table S8), consistent with the potential for a temperate lifestyle. In particular, vMAG_4615 encoded a CI-like repressor^51^, which may contribute to the maintenance of lysogeny and immunity against infection by related phages^15^ (Fig. 3). DNA methyltransferases were also identified in vMAG_1010, vMAG_4615, and vMAG_8907 (Table S8), potentially contributing to protection against host restriction systems^52^. No antibiotic resistance or virulence genes were identified in the four vMAGs.

These results provide complementary genomic evidence supporting the interaction of four candidate mycobacteriophages with NTM, with the strongest evidence for vMAG_4615 through its integration into the *M. arupense* genome. However, their actual host ranges, infectivity, and potential contributions to NTM survival or control remain to be experimentally validated.

### NTM concentrations showed distinct site-specific patterns and variable associations with putative mycobacteriophages

Bacterial and viral community composition differed significantly among sampling sites (PERMANOVA, R^2^ = 0.89, *p* = 0.003 for bacterial MAGs; R^2^ = 0.74, *p* = 0.004 for vOTUs). dbRDA showed associations between community composition and five physicochemical parameters (free ammonia, pH, temperature, electroconductivity, and nitrite) for both bacterial MAGs (*p* = 0.005) and vOTUs (*p* = 0.038) (Fig. S11). Site B was associated with free ammonia and nitrite, whereas Site A was associated with electroconductivity. However, variance partitioning showed that most of the explained variation was shared between physicochemical parameters and sample site (adjusted R^2^ = 0.72 for bacterial MAGs and 0.42 for vOTUs), with little variation uniquely explained by the physicochemical parameters (adjusted R^2^ = 0.00 and 0.09, respectively). Given the limited sample size and substantial overlap between physicochemical parameters and sampling site, their independent contributions to community composition could not be resolved. None of the measured parameters were significantly correlated with total NTM concentrations (Kendall, *p* = 0.176–1.000).

NTM concentrations were compared with the broader characteristics of the bacterial and viral communities (Table S9). Site E had the highest bacterial and viral alpha diversity (Shannon; Kruskal–Wallis, *p* = 0.027 for both; Fig. 4A and Fig. S12) and viral-to-bacterial ratio (*p* = 0.027; Fig. 1C). In contrast, Site B had the lowest values for these metrics. Total bacterial MAG and vOTU concentrations did not differ significantly among sites (Kruskal–Wallis, *p* = 0.67 and *p* = 0.25, respectively; Fig. S13). Mean NTM concentrations were highest at Site B (6.7 × 10^5^ ± 4.2 × 10^5^ copies/L), followed by Site A (4.7 × 10^5^ ± 2.1 × 10^5^ copies/L) and Site E (7.0 × 10^4^ ± 3.7 × 10^4^ copies/L; Fig. 4B), driven by strong differences between Sites A and E compared to Site B on the first two days (Days 1 and 3) of sampling. However, the NTM concentrations did not differ significantly among sites (Kruskal–Wallis, *p* = 0.061), as the NTM concentrations converged on Day 7. Mean NTM concentrations showed the opposite site-level ranking (B > A > E) to bacterial and viral Shannon diversity and the viral-to-bacterial ratio (E > A > B), though more samples are needed to confirm this.

**Figure 4.**
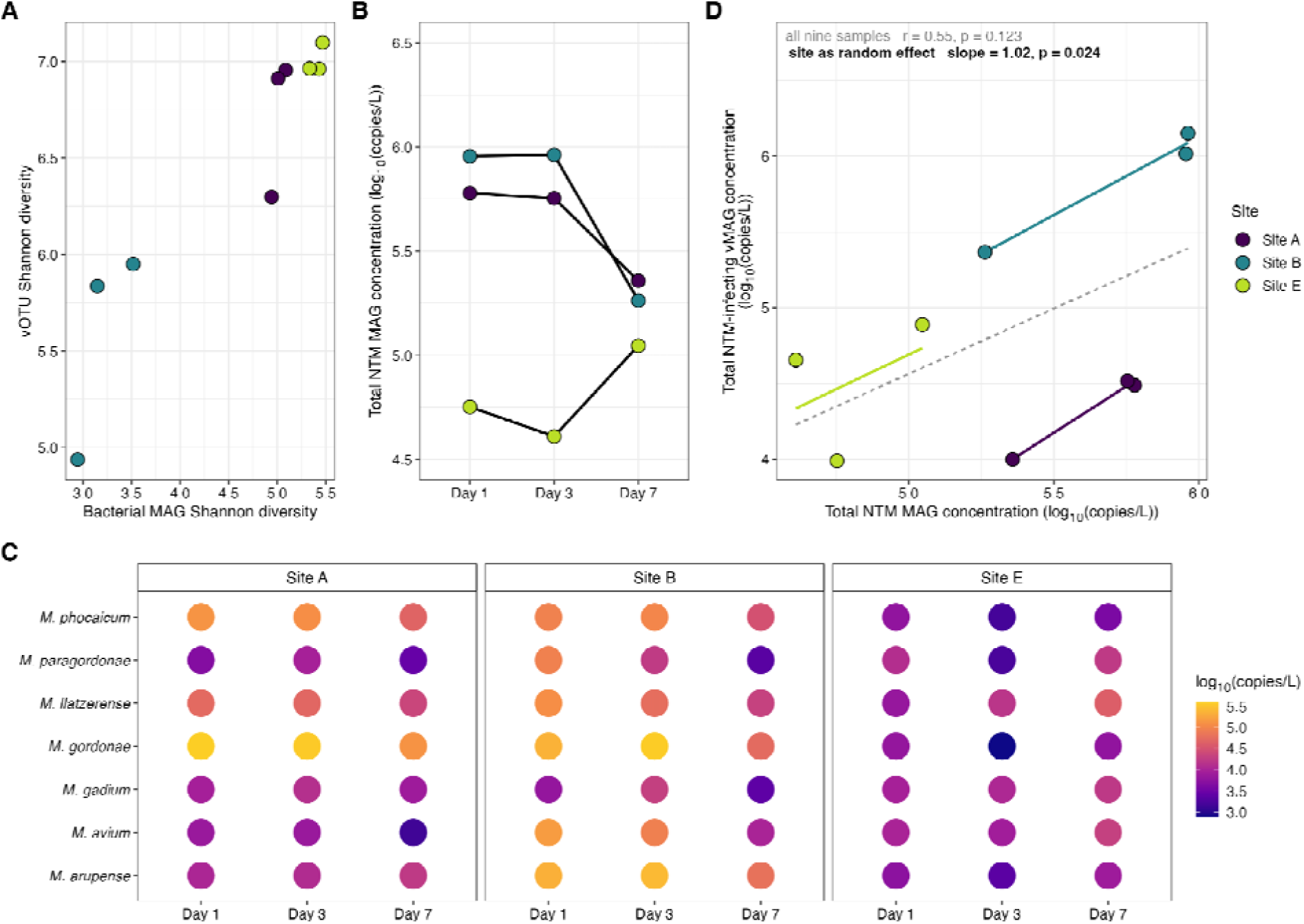
Site-specific patterns in bacterial, viral, and NTM MAG communities. (A) Shannon indices of bacterial and viral communities across nine samples. (B) Total NTM MAG concentrations (log_10_-transformed copies/L) across sampling days, grouped by site. (C) Total concentration of the four vMAGs assigned by iPHoP to *Mycobacterium* relative to total NTM MAG concentration across the nine samples (both log_10_-transformed copies/L). All nine sample were first analyzed using Pearson’s r (p-value reported), and a linear mixed-effects model wa fitted with site as a random effect (slope and p-value reported). (D) Bubble plot showing the concentrations of individual NTM MAGs across sites and sample days, with color representing log_10_-transformed copies/L.

While the relative ranking of NTM concentrations across the three sites matched prior surveillance data^26^ (Table S10), this pattern is not detectable with relative abundances. Using RPKM, Site A had a higher relative abundance of NTM than Site B (the total NTM MAGs averaged 1.55 RPKM at Site A, 0.81 RPKM at Site B, and 0.13 RPKM at Site E). Site B was previously characterized as having lower water use than Sites A and E^26^, although water use was not directly measured in the present study. The consistently elevated NTM concentration at Site B across years may therefore partially reflect lower local water use and associated stagnation, which could create conditions favorable to NTM persistence without producing a corresponding increase in overall microbial diversity.

NTM site patterns were not uniform across individual MAGs (Fig. 4D; Table S11). *M. arupense* had its highest mean concentration at Site B (1.81 × 10^5^ ± 1.04 × 10^5^ copies/L), whereas *M. gordonae* and *M. phocaicum* were most abundant at Site A (2.91 × 10^5^ ± 1.40 × 10^5^ and 9.88 × 10^4^ ± 4.59 × 10^4^ copies/L, respectively). All three occurred at substantially lower concentrations at Site E. However, none of the seven NTM MAGs remained significantly different among sites after Benjamini–Hochberg correction of the Kruskal–Wallis tests.

As an exploratory analysis, we also tested for potential relationships with amoebal hosts, because Megaviricetes, a group of giant viruses frequently associated with amoebae^53^, was the second most abundant viral class, and free-living amoebae can harbor NTM in drinking water^6^. Neither total eukaryotic nor *Acanthamoeba*-associated contig abundance was significantly correlated with total NTM concentration (Kendall, τ = −0.22, *p* = 0.477 and τ = −0.11, *p* = 0.761, respectively; Fig. S14). However, eukaryotic contigs accounted for only 0.2% of all contigs, and the workflow was not optimized for eukaryotic recovery^54^, which limits the interpretation of these negative associations.

We next examined whether NTM concentrations were associated with those of the four putative mycobacteriophages discussed above. The combined concentration of the four vMAGs was positively associated with total NTM concentration with site as a random effect (Fig. 4C; linear mixed-effects, slope = 1.02, *p* = 0.024). In contrast, the overall correlation across all nine samples was not significant (Pearson, r = 0.55, *p* = 0.12). The total concentration of the four vMAGs was higher at Site B compared to other sites (Wilcoxon rank-sum, *p* = 0.012), consistent with the pattern observed for the total NTM MAGs (Fig. 4B) and distinct from the overall pattern of the vOTUs, which did not vary across sites (Fig. S15). When examined individually, all four vMAGs showed positive associations with total NTM concentration, with significant associations observed for vMAG_8907 (linear mixed-effects, *p* = 0.005) and vMAG_8917 (linear mixed-effects, *p* = 0.017; Fig. S16). As iPHoP prediction is more reliable at genus-than species-level^55^, we examined whether concentration correlations for each vMAG–NTM MAG pair supported the predicted host associations. However, the observed correlations did not consistently align with the species-level host associations predicted by iPHoP (Fig. S17). For example, vMAG_8907 was positively associated with both *M. llatzerense*, a predicted host (linear mixed-effects, *p* = 0.011), and *M. avium*, which was not predicted by iPHoP (linear mixed-effects, *p* = 0.004). Conversely, vMAG_1010 showed no significant correlation with its predicted host, *M. llatzerense* (linear mixed-effects, *p* = 0.50). vMAG_4615, a predicted prophage of *M. arupense*, showed a significant positive correlation with *M. arupense* concentration (linear mixed-effects, *p* = 0.034). It also had a prophage-to-host copy number ratio of 0.90 ± 0.02, consistent with an integrated prophage without evidence of substantial lytic amplification during sampling. These patterns provide further support for associations between the four vMAGs and NTM, although they do not clearly distinguish their potential hosts at the species level.

### NTM MAGs encode a steroid degradation pathway largely absent from co-occurring Actinomycetota

Functional potential of the Actinomycetota MAGs in this system was compared to assess how NTM gene content differed from that of other species within the same phylum. The seven NTM MAGs and the eight other Actinomycetota MAGs recovered in this study (Table S6) yielded 2,216 and 2,333 distinct KEGG Orthology IDs (KOs), of which 1,558 were shared. The NTM MAGs had larger genomes than the other Actinomycetota (5.8 ± 1.4 vs. 3.6 ± 1.5 Mb; Wilcoxon rank-sum, *p* = 0.01) despite comparable genome completeness (86.0 ± 18.2 vs. 86.4 ± 9.4%; Wilcoxon rank-sum, *p* = 0.46; Table S6). However, average coding density was comparable between NTM and other Actinomycetota (1,037 ± 122 vs. 1,039 ± 52 predicted protein-coding genes per Mb; Wilcoxon rank-sum, p = 0.34).

The 20 KOs with the highest overall mean concentrations in NTM MAGs were primarily associated with transport and secretion, lipid metabolism, central metabolism, and stress response and regulation (Fig. S18; Table S12). Seventy-seven KOs were present in all seven NTM MAGs and absent from all eight Actinomycetota MAGs (Table S13): 14 in steroid degradation (discussed below), 13 in envelope glycolipid and mycolic acid synthesis, and 5 in the ESX (type VII secretion) system. These lipids contribute to envelope stability and reduced permeability and have been associated with antimicrobial resistance and pathogenicity^56–59^. ESX systems have been implicated in mycobacterial intracellular survival and pathogenicity^60–62^. Depending on the ESX subtype, they can contribute to phagosomal membrane disruption and intracellular survival in both amoebae and macrophages^62,63^, modulate host immune responses through PE/PPE protein secretion^63^, and mediate metal acquisition and homeostasis^17^. Notably, the enrichment of ESX-related genes in NTM MAGs may reflect their association with plasmids rather than their chromosomal gene content, as discussed in the following section.

A screen of chlorine- and oxidative-stress-related functions detected 39 distinct KOs across the seven NTM MAGs (Table S14), including the thioredoxin and peroxiredoxin system, glutaredoxins, mycothiol synthase, peptide-methionine (R)-S-oxide reductase (*msrB*), and the OxyR and SoxR regulators. While NTM had more distinct KOs (31 ± 4 vs. 24 ± 4; Wilcoxon rank-sum, *p* = 0.009) and more genes (54 ± 10 vs. 32 ± 6; Wilcoxon rank-sum, *p* = 0.002), the other Actinomycetota had a similar number of distinct KOs per Mb (5.6 ± 0.9 vs. 7.2 ± 2.1 per Mb; Wilcoxon rank-sum, *p* = 0.12) and the two groups carried the same number of genes per Mb (9.5 ± 1.7 vs. 9.5 ± 2.5 per Mb; Wilcoxon rank-sum, *p* = 0.87). In addition, the NTM MAGs carried an average of 2.9 × 10^6^ ± 2.5 × 10^6^ stress gene copies/L against 3.1 × 10^6^ ± 2.3 × 10^6^ in the eight other Actinomycetota MAGs (paired Wilcoxon signed-rank, *p* = 0.65), showing that NTM encoded no greater stress mitigation-related gene concentrations than other Actinomycetota in the same chloraminated water. No antimicrobial resistance genes were identified using AMRFinderPlus.

Among the 45 KEGG level-B categories, KOs related to xenobiotic biodegradation and metabolism showed the clearest difference: an average of 8.2 ± 1.8 KOs per Mb in NTM, compared with 2.6 ± 1.8 KOs per Mb in the other Actinomycetota (Wilcoxon rank-sum, Benjamini–Hochberg q = 0.002). The steroid degradation pathway explained most of this difference (6.0 ± 1.4 KOs per Mb in NTM vs. 0.8 ± 0.5 in the other Actinomycetota; Fig. S19; Wilcoxon rank-sum; Benjamini–Hochberg q = 0.003; Fig. 5). Additionally, the NTM MAGs carried substantially more steroid degradation gene copies (3.5 × 10^6^ ± 3.0 × 10^6^ copies/L vs. 2.1 × 10^5^ ± 1.5 × 10^5^ in the eight other Actinomycetota MAGs; Wilcoxon signed-rank, q = 0.026; Fig. S20). Each NTM MAG carried 31 to 36 of the 37 detected steroid degradation KOs, compared with 1 to 11 in the other Actinomycetota, and 14 KOs covering all four steps of the pathway were found in all seven NTM MAGs but none of the eight Actinomycetota MAGs (Fig. 5; Table S13). These results indicate that NTM disproportionately contained steroid degradation genes among Actinomycetota in this system.

**Figure 5.**
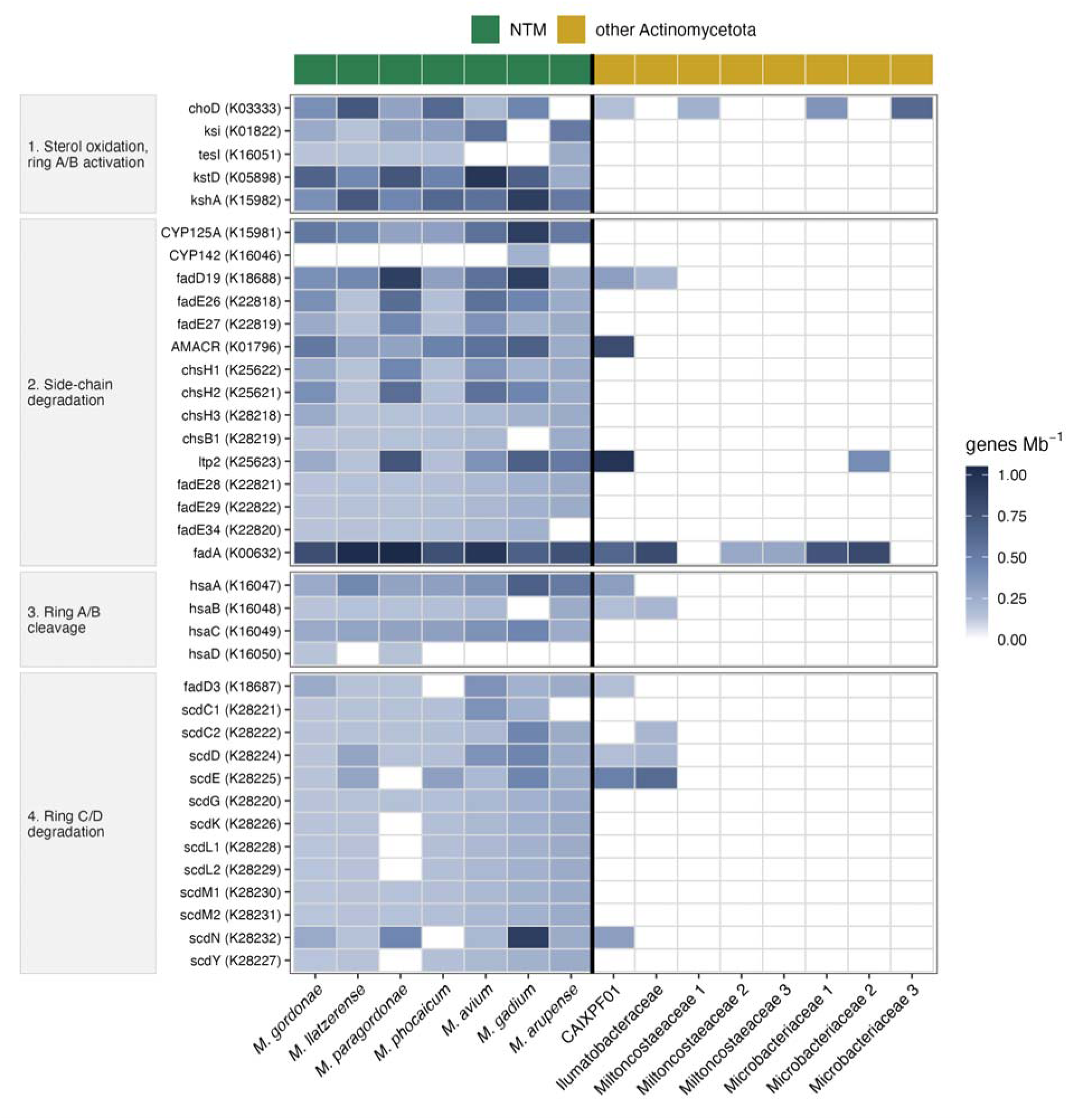
Steroid degradation genes of NTM and co-occurring Actinomycetota MAGs. The 37 KEGG Orthology IDs of the steroid degradation pathway detected in this study are shown for the seven NTM MAGs and the eight other Actinomycetota MAGs recovered here, arranged in four blocks following the mycobacterial cholesterol catabolism route: sterol oxidation and ring A/B activation, side-chain degradation, ring A/B cleavage, and ring C/D degradation. Shading indicates the gene copy number per Mb for each MAG, with white indicating zero.

Steroid catabolism in mycobacteria is associated with cell envelope maintenance^64^, redox homeostasis^65^, and metabolic flexibility^66^, suggesting potential roles in adaptation to environmental stress. In *M. tuberculosis*, cholesterol utilization is required for persistence during chronic infection and growth within activated macrophages^67^. Moreover, steroid metabolism provides NTM with a competitive advantage: not only can NTM grow on cholesterol as a sole carbon source, but cholesterol amendment of sewage influent enriched mycobacteria 4.7-fold within seven days relative to the background bacterial community^68^. Although sterols are generally present at low concentrations in drinking water, steroid degradation may contribute to NTM survival within environmental hosts. Free-living amoebae have sterol-containing membranes and could potentially provide a source of sterols for intracellular NTM^69^. However, whether NTM utilize these sterols or whether steroid degradation contributes to their survival in drinking water remains unknown. Additionally, there is no evidence that these genes are being transferred horizontally. While 30 putative horizontal gene transfer events were identified between NTM and co-occurring bacterial taxa (Table S15), none of these were related to steroid metabolism. Future studies are needed to determine whether NTM utilize environmental or host-derived sterols and whether steroid degradation contributes to their survival in drinking water or during human infection.

### NTM plasmids carry genes that may contribute to survival of NTM in drinking water

In total, 438 plasmid-associated contigs (Text S5) were detected in all seven NTM MAGs (median geNomad plasmid score 0.95; 295 scoring ≥0.9), representing a mean of 14.8 ± 5.3% of the host MAG length. The *M. paragordonae* MAG had the highest number of plasmid-associated contigs (146), followed by *M. gordonae* (75) and *M. gadium* (70) (Table S16).

Four plasmid-associated contigs matched three *Mycobacterium* reference plasmids in PLSDB^70^ at 97.8–99.8% nucleotide identity over 97.9–100% of the contig (Table S17). These included one contig in the *M. arupense* MAG matching a *Mycobacterium intracellulare* subsp. chimaera plasmid and one in the *M. paragordonae* MAG matching a *Mycobacterium kansasii* plasmid. Together, two contigs in the *M. phocaicum* MAG spanned non-overlapping parts of the plasmid, almost completely covering pGD25-3 from a clinical *M. abscessus* isolate. All three references are closed circular sequences, whereas the matching contigs were incomplete and had no terminal repeats. Each reference is classified as non-mobilizable yet encodes a mobility-associated gene (Table S17), either a MobF family relaxase or a type IV secretory system conjugative DNA transfer protein^71,72^. The detection of plasmids closely related to those associated with clinical NTM isolates suggests that drinking water and clinical NTM plasmid pools may overlap. While these data do not demonstrate direct transmission or identify specific virulence-associated transfer events, they suggest that drinking water could contribute to the maintenance or exchange of plasmid-associated traits relevant to NTM persistence, stress tolerance, or pathogenicity.

Contigs predicted as plasmids in NTM MAGs carried an average of 5.7 × 10^8^ gene copies/L, representing 20.2 ± 8.5% of total NTM gene copies (range, 9.3–37.4% across nine samples). No antimicrobial resistance genes were detected despite prior reports of such genes in *M. avium* and *M. intracellulare* plasmids^73^. Functional annotation identified 532 KOs in NTM plasmids. The 20 KOs with the highest mean absolute concentrations were dominated by plasmid partitioning and mobility, type VII secretion, metal efflux, and envelope lipid transport (Fig. S21).

We compared the proportion of annotated genes assigned to each KEGG level-C category between plasmid-associated and chromosomal contigs within each of the seven NTM MAGs (Table S18). Secretion-system genes made up 11.6% of the KO-annotated genes on plasmid-associated contigs compared with 1.7% of those on chromosomal contigs (Fisher’s exact test, Benjamini–Hochberg, q < 0.001). Type VII secretion accounted for most of these genes and spanned the complete set (Fig. 6): *EccA–E*, the mycosin protease *MycP*, the ESAT-6 substrate *EsxA*, and *EspA/E, EspG, EspI*, and *EspK*. A mycobacterial conjugative plasmid requires both type IV and type VII secretion for transfer^17,74^; while a type IV secretion cluster (*trbB*–*trbE*) was present on a plasmid-associated contig, no contig carried both systems. Beyond their potential role in plasmid transfer, the enrichment of ESX genes on plasmid-associated contigs suggests that plasmids may contribute to NTM functions involved in host interactions and intracellular survival, as discussed above. Metal-efflux-related genes comprised 5.3% of plasmid-associated annotations versus 0.5% of chromosomal annotations (Fisher’s exact test, Benjamini–Hochberg, q < 0.001) (Fig. 6), including the copper exporter *copA*, the Zn² /Cd² exporter *zntA*, and mercuric reductase *merA*. Copper and zinc/cadmium efflux functions further suggest a potential role for plasmids in metal homeostasis in NTM, with copper efflux potentially relevant to exposure from copper pipe surfaces at Sites B and E (Table S1). Plasmids can expand drinking water NTM’s pool of genes with functions beneficial to drinking water survival and human infection, but their expression and effects remain to be experimentally confirmed.

**Figure 6.**
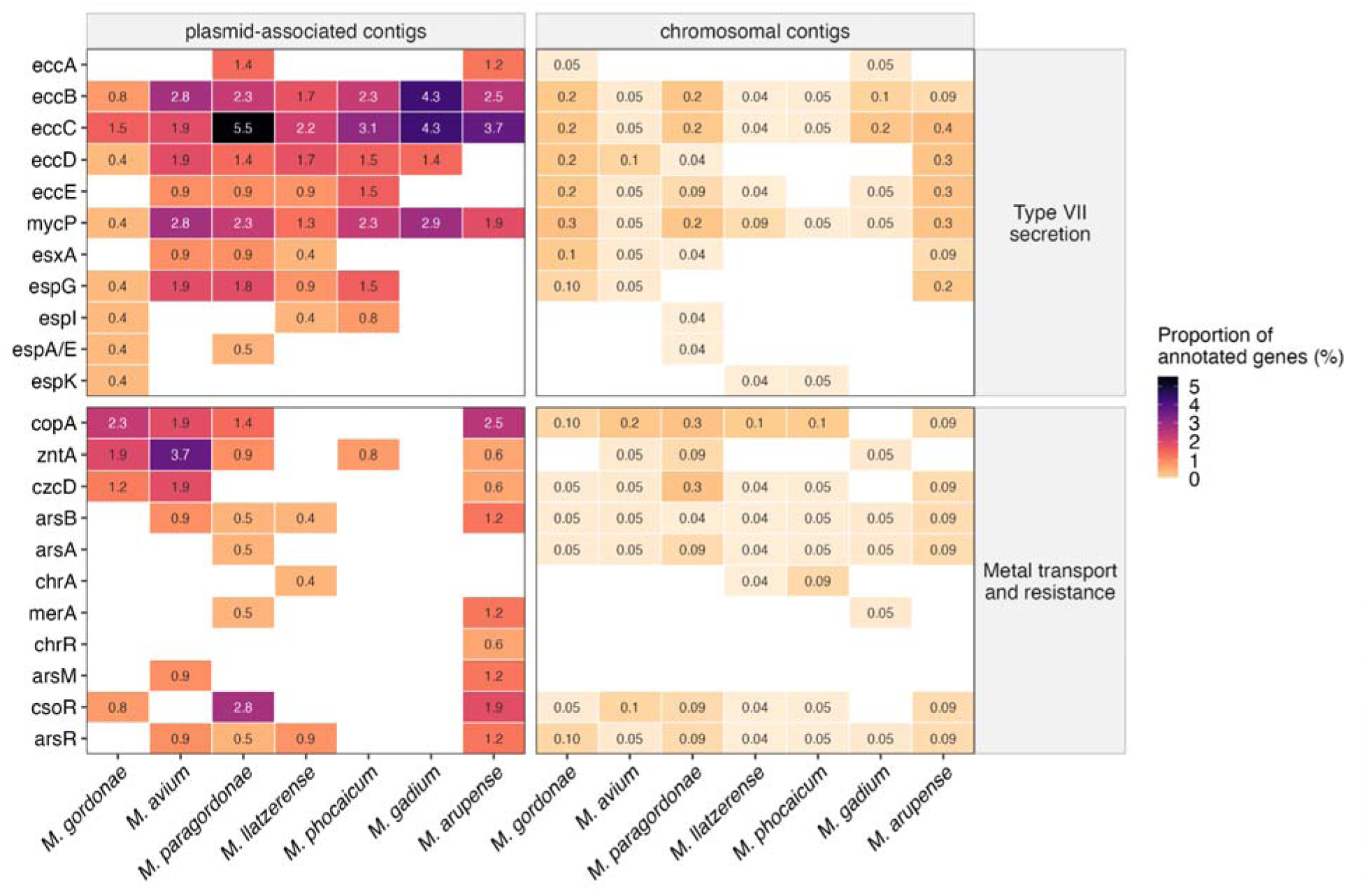
Metabolic potential of NTM-associated plasmids. Type VII secretion (ESX) and metal transport genes on plasmid-associated and chromosomal contigs of seven NTM MAGs. Each cell shows the percentage of KO-annotated genes assigned to the indicated gene within each MAG and genomic compartment. Percentages were calculated by combining all contigs in each compartment for each MAG. Chromosomal contigs were defined as those classified as neither plasmids nor proviruses. Blank cells indicate zero, and values below 0.1% are displayed to two decimal places.

### Future Directions

Genome-resolved quantitative metagenomics revealed site-specific NTM abundance patterns that contrasted with broader bacterial and viral community trends. Putative mycobacteriophag concentrations were positively correlated with total NTM concentrations within sites but not across the full sample set. We also showed that plasmids contain genes that may contribute to NTM survival in drinking water and human hosts. Future work should investigate whether these ecological and genomic patterns recur across drinking-water systems with different hydraulic conditions, plumbing materials, and disinfectant regimes. Metatranscriptomic and metaproteomic analyses could identify functions active in situ, while sequencing targeting eukaryotes combined with microscopy-based assays could elucidate interactions among viruses, amoebae, and NTM. We observed that bacteria, like NTM, with high GC content had more predicted phage-host interactions. Approaches establishing physical rather than computational phage–host linkage, together with isolation of candidate mycobacteriophages, will be needed to confirm host ranges, infection dynamics, and potential for biocontrol. As multiple NTM species are included on the draft CCL 6, these findings underscore the value of quantitative, genome-resolved surveillance for characterizing the gene pools of drinking water NTM and their associated phage- and plasmids.

## Data availability

All raw sequencing reads for this study have been deposited at NCBI under the bioproject number PRJNA1514785. The associated code for this project is available in the project’s GitHub repository [github.com/HegartyLab/DW_phage_plasmids_NTM].

## Acknowledgments

We would like to thank the Remaking Water Infrastructure Blue Sky team, particularly Tarik Quneibi, Matthew Vedrin, and Katherine Dowdell, as well as the Raskin, Hegarty, Duhaime, and Wigginton labs, for their technical and intellectual expertise in bioinformatics, experimental design and analyses, and manuscript preparation. We thank the staff of the City of Ann Arbor water treatment plant, as well as the building owners and staff, for their support of our sampling. This work was supported by the Blue Sky Initiative of the University of Michigan College of Engineering, as well as Dr. Hegarty’s CAREER Award (Award Number 2442755) and startup funds from the Case Western Reserve University Case School of Engineering. We gratefully acknowledge the American Chemical Society (ACS) Graduate Student Success Grant for supporting this research. This study was supported by the University of Michigan Advanced Genomics Core. During the preparation of this manuscript, the authors used ChatGPT and Claude for language editing and to improve clarity. The authors subsequently reviewed and revised the content and take full responsibility for the final manuscript.

