## Supplemental Methods and Figures for "Genome-resolved quantitative metagenomics of nontuberculous mycobacteria and their associated phages and plasmids in chloraminated drinking water"

### Supplemental text

### Text S1: Detailed short-read sequence processing, de novo co-assembly, binning, and taxonomy assignment.

**Pre-processing short-read sequences.** Short-read metagenomic sequence processing followed the workflow described by Vosloo et al. (2021)^1^. Adapter removal and quality filtering were performed using fastp v0.23.2 with the flags --qualified_quality_phred 20, --trim_poly_g, --trim_poly_x, and --length_required 20^2^. The UniVec Core database from NCBI (ftp://ftp.ncbi.nih.gov/pub/UniVec/) was subsequently used to identify potential vector-derived sequences using BWA-MEM v0.7.17^3^. SAMtools v1.9 was used for filtering, and reads were extracted using bedtools v2.30.0^4^. The resulting quality-filtered FASTQ files were analyzed using Nonpareil v3.53^5^ in kmer mode to estimate sequence coverage, diversity, and redundancy. The quality-filtered forward and reverse FASTQ files for each sample were then interleaved using Interleafq v1.1.0 (<https://github.com/quadram-institute-bioscience/interleafq>).

**Co-assembly of short-read sequences.** Before assembly, interleaved reads from all samples were pooled and co-assembled with metaSPAdes v4.0.0^6^ using k-mer sizes of 21, 33, 55, 77, 99, and 127. Contigs smaller than 1 kb were removed from the assemblies using seqtk (<https://github.com/lh3/seqtk>), followed by using the dedupe function of BBTools v39.03 to remove redundant contigs, including duplicate and contained contigs (<https://github.com/BioInfoTools/BBMap/blob/master/sh/dedupe.sh>). QUAST v5.2.0^7^ was utilized to evaluate the quality of the processed assemblies using default parameters. Quality-filtered reads were mapped back to the co-assembly using BWA-MEM v0.7.17^3^. SAM files were filtered using the view function of SAMtools v1.9 to retain only properly mapped reads using parameters -hbS and -F4^4^. BAM files were sorted, and mapping rates were calculated using CoverM v0.6.1^8^ to assess contig coverage depth with the flag set to count, yielding 97.75 ± 0.49%.

**Binning of short-read sequences.** Contigs larger than 1 kb were used for binning in Anvi’o v8.0^9^. Initial bins were generated with MetaBAT v2.15^10^, yielding 895 bins. Bin quality was evaluated with CheckM v1.2.1^11^. Mapping rates were determined using CoverM v0.6.1^8^ to assess bin coverage depth, following the alignment of quality-filtered reads to the assembly with BWA-MEM v0.7.17^3^. SAM files were filtered with SAMtools v1.9 using the parameters -hbS and -F4 to retain only properly mapped reads^4^. The resulting BAM files were then sorted, yielding an average mapping rate of 97.28 ± 0.74%. Bins with completeness over 50% and redundancy below 10% were manually refined using anvi-refine from Anvi’o v8.0^9^ , yielding 522 medium-quality MAGs^12^. After refinement, reads were remapped, and coverage recalculated as described above. The average read mapping rate to the refined MAG set was 87.00 ± 3.38%.

**Taxonomy and phylogeny of short-read sequences.** The Contig Annotation Tool (CAT/BAT) v6.01 was used to perform taxonomic assignment of 522 MAGs from superkingdom to species level^13^ (Table S19). Based on CAT/BAT, seven MAGs were assigned to the family Mycobacteriaceae; at genus level, three of these were identified as *Mycobacterium*, one as *Mycolicibacter* and one as *Mycolicibacterium*, while two received no genus-level assignment. In addition, taxonomy was assigned using GTDB-Tk v2.4.0 with the "classify_wf" flag and the GTDB reference database release 220^14^. Based on the combined CAT/BAT and GTDB-Tk results, seven MAGs were classified as NTM MAGs, with different species-level identities. The ANI of the *M. avium* MAG was calculated using fastANI v1.34^15^. Recent phylogenomic analyses^16^ have proposed division of the genus *Mycobacterium* into multiple genera, including *Mycobacteroides*, *Mycolicibacterium*, *Mycolicibacter*, and *Mycolicibacillus*, based on genome-scale evidence. In this study, the traditional genus designation *Mycobacterium* was retained for consistency with much of the current clinical and environmental NTM literature and to maintain terminological continuity. GToTree v1.8.6^17^ was used to construct a phylogenomic tree of NTM MAGs based on concatenated single-copy marker genes. Maximum-likelihood tree inference was performed using FastTree as implemented in GToTree. Genomes that did not meet the default threshold of ≥50% marker gene presence were excluded; as a result, two of the seven NTM MAGs were not included in the final tree. The tree was rooted using *Yersinia pestis* as a closely related outgroup and visualized using Interactive Tree of Life (iTOL; v6)^18^. Correlations between the MAG sizes of these seven NTM MAGs and their corresponding reference genome sizes (Fig. S1) were used to assess whether genome recovery was sufficient to support downstream analysis, including quantification of MAG concentrations, identification of plasmid and prophage contigs, and assessment of functional potential.

**Text S2: Full-flush samples.**

After flushing until the distribution system water was reached, full-flush samples were collected at Site A once temperature, pH, and oxidation–reduction potential (ORP) had stabilized, as previously described^19,20^. A 10 L water sample was collected at each time point and filtered through a Sterivex 0.22 µm filter for long-read sequencing. After filtration and elution, the three full-flush samples were stored at -80 °C.

**Text S3: Long-read sequencing and assembly.**

Long-read sequencing was performed by the University of Michigan Advanced Genomics Core. Initial QC was performed to ensure that samples met Oxford Nanopore Technologies (ONT) requirements for library preparation, using Qubit for DNA quantification, NanoDrop for purity assessment (260/280 and 260/230 ratios), and Agilent TapeStation for fragment size estimation. Following QC, samples were bead-cleaned using Ampure XP beads (1.8× ratio) to achieve the NanoDrop purity levels specified by ONT. Library preparation was then performed using the Native Barcoding Kit 24 v14, with specific barcodes assigned to each sample to enable multiplexing. Samples were then normalized to equal mass, barcoded, and pooled. The pooled library was then sequenced on an R10.4.1 MinION flow cell with the super-accurate basecalling model using a GridION instrument. A total of two library loads were performed, with a nuclease wash between the loads, to maximize sequencing throughput.

LongQC v1.2.0c^21^ and FastQC v0.12.1 (<https://github.com/s-andrews/FastQC>) were used to assess the long-read quality. Reads were processed with fastplong v0.2.2 (<https://github.com/OpenGene/fastplong>) to trim the first 10 bp and remove reads exceeding 30,000 bp for quality filtering. Viral contigs were assembled from the quality-filtered reads using ViralFlye v0.2^22^. Assemblies were polished using Racon v1.5.0^23^ with four iterative rounds, which used minimap2 v2.29-r1283^24^ to align reads to the assembly. Polished assemblies were further refined using Medaka v2.1.0 (<https://github.com/nanoporetech/medaka>). QUAST v5.0.2^7^ was used to evaluate the quality of the assembly and polished assembly.

**Text S4: Viral sequence identification and host prediction.**

**Viral sequence identification**. geNomad (v1.8.1 for short reads and v1.8.0 for long reads) was used to identify viral contigs and assign taxonomy, with default settings^25^. Non-redundant vOTUs were then defined by clustering contigs at 95% ANI across at least 85% of their length, consistent with species-level viral clusters^26^. For each cluster, the longest contig greater than 3000 bp was retained as the representative vOTU sequence.

The vOTUs were compared to the VHIP database of experimentally confirmed viral–host infection pairs^27^ and to the complete Actinophage database^28^ using BLAST (BLAST+ v2.16.0)^29^. Hits were retained when the aligned length was at least 1 kb; nucleotide identity and aligned length are reported in Table S7. VIBRANT v1.2.1^30^ was used to identify vOTUs with markers of lytic and lysogenic lifestyles. Viral tRNAs were identified using ARAGORN v1.2.41^31^ and then BLASTed against the MAGs to find exact matches. The Proksee web server was used for visualizing a select set of vOTUs^32^.

**Viral host prediction.** Virus-host infection potential was evaluated using VHIP^27^ and iPHoP^33^. VHIP assigns scores (0 to 1) to vOTUs based on the likelihood of infecting a given MAG. Based on these scores, interactions were classified as predicted non-interacting (interaction = 0, score < 0.99) or predicted interacting (interaction = 1, score ≥ 0.99). The default VHIP score cutoff was 0.5, but we increased it to 0.99. Viral–host pairs shared across sites were used to compare predicted interaction networks across sites and days. Due to VHIP requirements, only vOTUs with a completeness score greater than 75% (vMAGs) from CheckV v1.0.3 were used to predict virus–host infection networks. These vOTUs were classified as vMAGs^34^. The extensive predicted host sharing persisted under more stringent genome completeness criteria: increasing the viral completeness threshold from 75% to 90% increased rather than decreased the proportion of vMAGs predicted to interact with all seven NTM MAGs (Fig. S22). To test whether the extensive predicted host sharing among the NTM MAGs reflected their narrow, high GC range, the observed proportion of vMAGs predicted to interact with all seven NTM MAGs was compared with the same proportion for comparison sets of seven bacterial MAGs. Four sets were drawn at random 2,000 times each, from all 522 MAGs, from within a single phylum, from the 102 non-NTM MAGs whose GC content falls within the NTM range of 65.0–69.2%, and from the 84 Pseudomonadota MAGs within that range. A fifth comparison used all eight possible sets of seven drawn from the eight non-NTM Actinomycetota MAGs, which is an exhaustive enumeration rather than a random draw. Medians and the 2.5th and 97.5th percentiles across the draws are reported. Separately, the GC difference between each virus and host was calculated as the vMAG GC content minus the MAG GC content, and predictions with a difference below −20%, a range sparsely represented in VHIP's training pairs, were excluded as a sensitivity analysis. iPHoP assigns a confidence score (0-100) to each phage based on the likelihood that it will infect a bacterium in its database. Following iPHoP’s recommendations^33^, infection was predicted for those with a confidence score greater than 90. We ran iPHoP against both its default database and a custom database incorporating the seven NTM MAGs recovered in this study.

**Text S5: Plasmid and prophage identification.**

Plasmid and prophage contigs were identified from MAGs using geNomad v1.8.1^25^ from the assembled contigs ≥ 1 kb, independently of genome binning. The resulting contigs were then aligned against the NTM MAGs using BLASTn v2.17.0^35^. For each candidate plasmid or prophage, query coverage was calculated as the alignment length divided by the total contig length. Only sequences with 100% query coverage (i.e., the entire contig aligning back to the NTM MAGs) were retained for downstream analysis and categorized as NTM plasmids and prophages. Plasmids were then screened against the plasmid sequence database PLSDB (<https://ccb-microbe.cs.uni-saarland.de/plsdb2025>)^36^ and each contig with a reference match was then realigned to that reference with BLASTn to obtain nucleotide identity and alignment coverage. Plasmid and prophage absolute abundances were obtained as their read-coverage ratio to the host chromosome, calculated using featureCounts from the Subread package^37^, multiplied by the absolute abundance of the host MAG.

**Text S6: Metabolic potential analysis.**

Predicted protein sequences from NTM MAGs and their associated plasmid and prophage regions were annotated using GhostKOALA based on KEGG Orthology^38^. Eight other Actinomycetota MAGs recovered in this study were also annotated using the same GhostKOALA workflow. Chlorine- and oxidative-stress genes were screened against a list of 154 KOs compiled from published chlorine-stress mitigation and oxidative-stress gene sets. Antimicrobial resistance genes were identified using AMRFinderPlus v4.0.23^39^.

For each of the seven NTM MAGs, contigs predicted as plasmids were combined into a plasmid-associated group, contigs predicted as prophages were combined into a prophage-associated group, while contigs classified as neither plasmids nor prophages were assigned to the chromosomal group. For each MAG and genomic compartment, the proportion of genes assigned to a given functional category was calculated by dividing the number of KO-annotated genes assigned to that category by the total number of KO-annotated genes in the corresponding compartment. Categories absent from a compartment were assigned a proportion of zero. Gene proportion of MAGs was compared between the seven NTM MAGs and the eight other Actinomycetota MAGs by dividing the number of distinct KOs assigned to a KEGG category by that MAG's assembly length (Mb). Comparisons used two-sided Wilcoxon rank-sum tests, with Benjamini–Hochberg correction applied. Average coding density, expressed as predicted protein-coding genes per Mb of assembly, and assembly length were compared between the two groups of MAGs with the same test, without correction. Gene proportions of plasmids were compared between plasmid-associated and chromosomal contigs with a two-sided Fisher's exact test on KO-annotated gene counts, run separately within each of the seven MAGs against its own chromosome, with Benjamini–Hochberg correction across the seven tests. Percentages are means across the seven MAGs. Categories or pathways absent from a MAG were assigned zero before testing. Corrected values are reported as q.

Gene-level absolute abundances were estimated from gene copy number. Each predicted gene was assigned the QuantMeta-derived absolute abundance (copies/L) of the sequence carrying it, so that a gene present in a single copy is reported at the absolute abundance of its replicon. For each MAG, plasmid, or prophage, the absolute abundances of genes assigned to the same KO were summed within each sample to obtain KO-level concentrations. KO-level concentrations were then averaged across the nine samples. Where a concentration is reported for a group of genomes, the value given is the mean across that group's MAGs of each MAG's gene concentration. Gene concentrations were compared between the two groups with a two-sided Wilcoxon signed-rank test on log_10_-transformed values, paired by sample.

For horizontal gene transfer, MetaCHIP v1.10.10^40^ was used to identify genes potentially donated to or received by NTM MAGs from other bacterial MAGs. Putatively transferred genes were further analyzed using BLASTp^29^ (BLAST+ v2.17.0;<https://blast.ncbi.nlm.nih.gov/Blast.cgi?PAGE=Proteins>) with query coverage > 90%.

Genes in the four candidate vMAGs and the five prophage regions were called from the geNomad-extracted regions and annotated from three sources: KEGG Orthology from GhostKOALA^38^; KO, Pfam, and VOG assignments from VIBRANT^30^, both joined on the gene index within the region; and the geNomad v1.8.1 protein description and virus-hallmark flag^25^, joined on coordinate overlap because geNomad numbers genes across the whole contig. Each gene was assigned one functional category by keyword matching against the combined annotation text, with sources taken in a fixed order so that a general geNomad description does not override a specific assignment, and genes whose only match was to a hypothetical or unknown protein were left unannotated. For each vMAG, the proportion of annotated genes whose best VOG hit named mycobacteriophage L5 or D29 was compared between the four candidate vMAGs and the other 316 annotated vMAGs with a two-sided Wilcoxon rank-sum test.

**Text S7: Statistical analysis.**

All statistical analyses and visualizations were conducted in RStudio (2023.09.1+494) with R (v4.4.0) unless otherwise noted. Figures were generated using ggplot2 (v3.5.2) unless otherwise noted. Heatmaps were created using pheatmap (v1.0.13), with dendrograms drawn by complete-linkage hierarchical clustering (hclust, stats v4.4.0) where noted. All tests were two-sided, and statistical significance was defined as p < 0.05. Benjamini–Hochberg corrected values are reported as q.

Alpha diversity of the bacterial MAG and vOTU communities was calculated from per-sample read counts using the Shannon and Simpson indices with vegan (v2.6-10).

Pearson correlation analysis was used to examine linear relationships between total bacterial MAG and vOTU concentrations (log10-transformed), between the number of predicted NTM hosts and the total number of bacterial hosts for each vMAG, between the combined concentration of the four candidate NTM-associated vMAGs and total NTM concentration (log10-transformed + 1), and between individual candidate vMAGs and their corresponding NTM MAGs (log10-transformed + 1). To evaluate covariation within sites, linear mixed-effects models were fitted with site as a random intercept using lme4 (v1.1-35)^41^ and lmerTest (v3.1-3)^42^, in the form log10(y + 1) ~ log10(x + 1) + (1|site). This model was used for the relationship between total vOTU and total bacterial MAG concentration, between the combined concentration of the four candidate NTM-associated vMAGs and total NTM concentration, between each of those four vMAGs individually and total NTM concentration, and for each of the 26 VHIP-predicted vMAG–NTM MAG pairs. Fixed-effect slopes and their p-values are reported, and models whose random-effect variance was estimated at zero are identified as singular fits.

Differences among sites were tested using the Kruskal–Wallis test for bacterial and viral Shannon and Simpson diversity, the viral-to-bacterial ratio, total bacterial MAG concentration, total vOTU concentration, total NTM concentration, and the concentration of each of the seven NTM MAGs. With three sites and three samples per site, the smallest p-value attainable by this test is 0.027. Benjamini–Hochberg correction was applied within three families of tests: the seven site comparisons of individual NTM MAGs, the 26 linear mixed-effects models of individual vMAG–NTM MAG pairs, and the comparisons of functional gene content between NTM and other Actinomycetota MAGs (Text S6). Comparisons of gene counts, gene density, genome size, coding density, and gene concentrations between MAG groups used two-sided Wilcoxon rank-sum or signed-rank tests, and comparisons of gene proportions between plasmid-associated and chromosomal contigs used two-sided Fisher's exact tests; these are described in Text S6.

Kendall's rank correlation was used to evaluate associations between total NTM concentration and physicochemical parameters, total eukaryotic sequence abundance, and Acanthamoeba-associated sequence abundance. For each sample, eukaryotic sequence abundance was calculated by summing contig length multiplied by mapped-read count across contigs classified as Eukaryota by the Contig Annotation Tool (CAT v6.01, lineage score fraction f = 0.5); Acanthamoeba-associated sequence abundance was calculated in the same manner using contigs assigned to the genus Acanthamoeba. These variables were analyzed untransformed.

Community composition was analyzed from per-sample read-count matrices of bacterial MAGs and vOTUs, Hellinger-transformed, using vegan (v2.6-10). Differences among sites were tested by PERMANOVA (adonis2). Distance-based redundancy analysis (dbRDA) was then used to relate community composition to physicochemical parameters. To reduce collinearity among environmental predictors prior to dbRDA, pairwise Pearson correlations were calculated for all measured water chemistry variables. Variables with absolute correlation coefficients greater than 0.8 were considered redundant and were excluded from the same model (Fig. S23). Based on this screening, free ammonia, pH, temperature, electrical conductivity, and nitrite were retained as the reduced predictor set. Overall model significance was assessed by a permutation test. Because nine samples and five predictors leave three residual degrees of freedom, variance partitioning (varpart) was used to separate the variation explained by the physicochemical parameters from the variation explained by sampling site, and adjusted R² values are reported for the shared and unique fractions.

### Supplemental figures

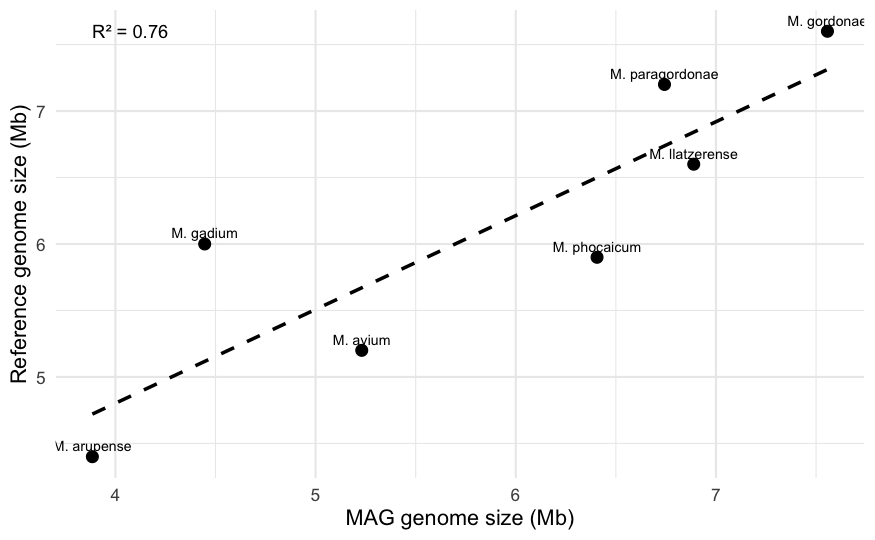

**Figure S1**. Relationship between the assembled sizes of NTM MAGs and the genome sizes of their corresponding reference species. Each point represents one MAG (labeled by species), and the dashed line indicates the linear regression fit (R^2^ shown).

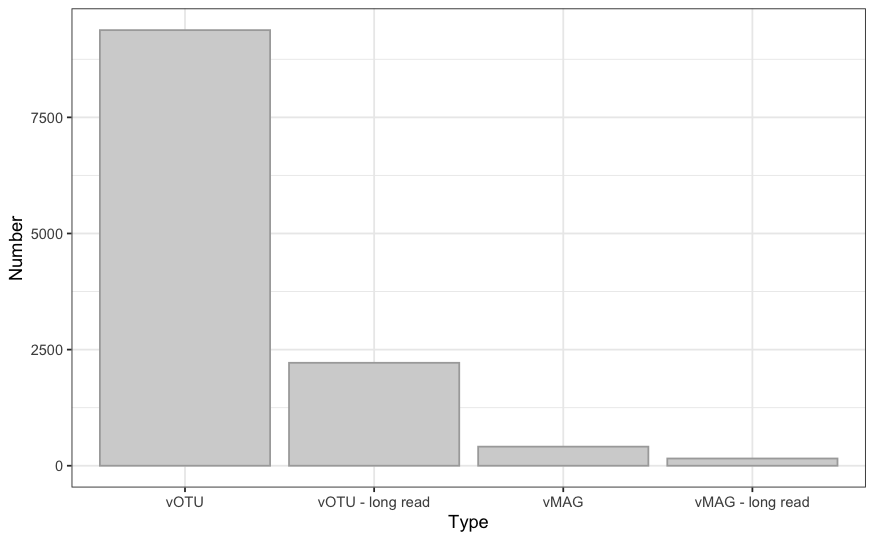

**Figure S2.** vOTU counts categorized by assembly source and completeness. From left to right: all vOTUs detected in short-read data (vOTU), vOTUs detected in long-read contigs (vOTU - long read), vMAGs (completeness >75%) recovered from short-read data (vMAG), and vMAGs detected in the long-read contigs (vMAG - long read).

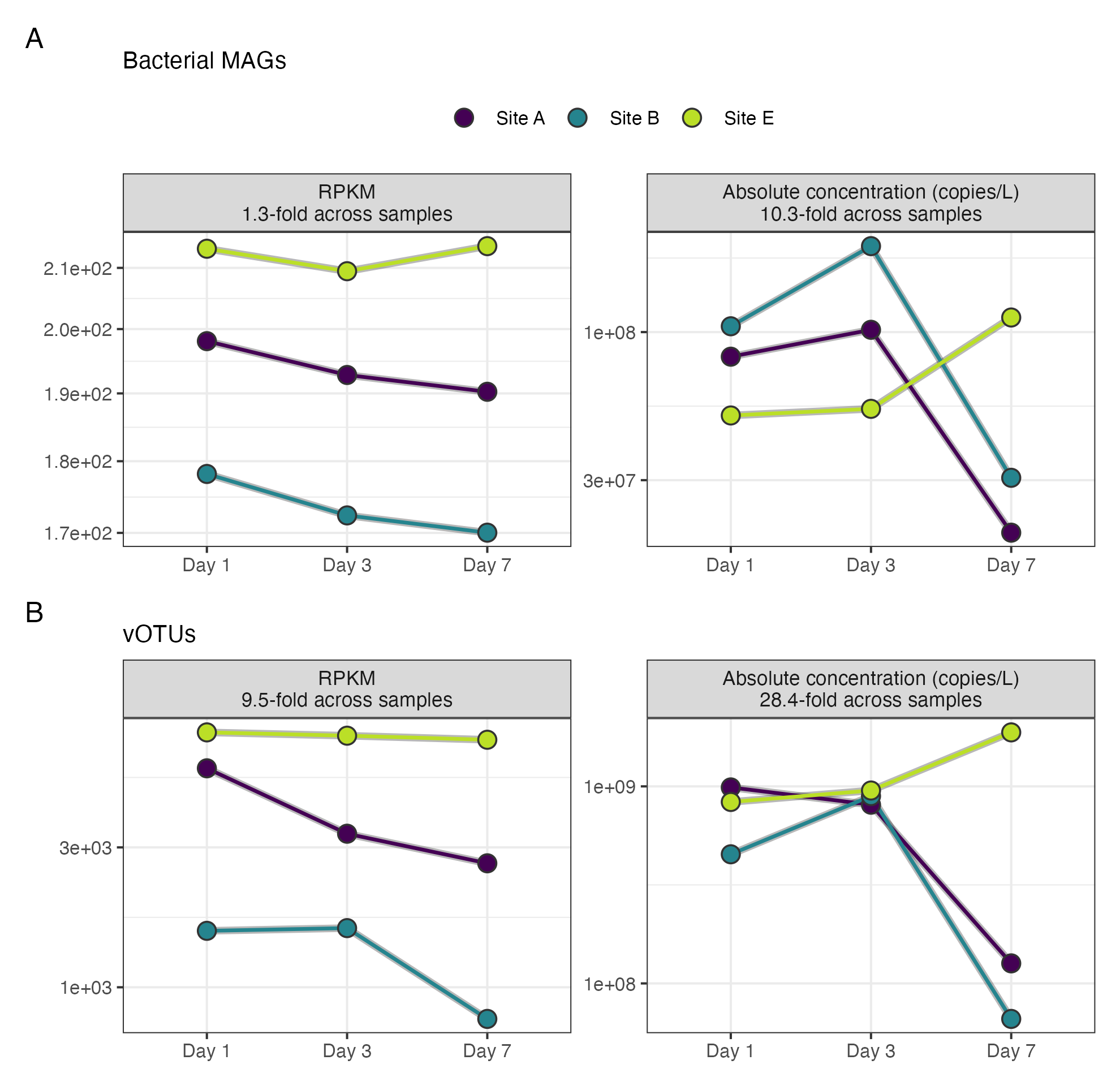

**Figure S3.** Sequencing depth does not track absolute abundance. Per-sample totals across the nine first-draw samples, colored by site, with log-scaled y-axis. (A) Bacterial MAGs: RPKM values mapped to the MAGs span 1.3-fold across samples, whereas total absolute concentration spans 10.3-fold. (B) vOTUs: RPKM values span 9.5-fold and total absolute concentration 28.4-fold.

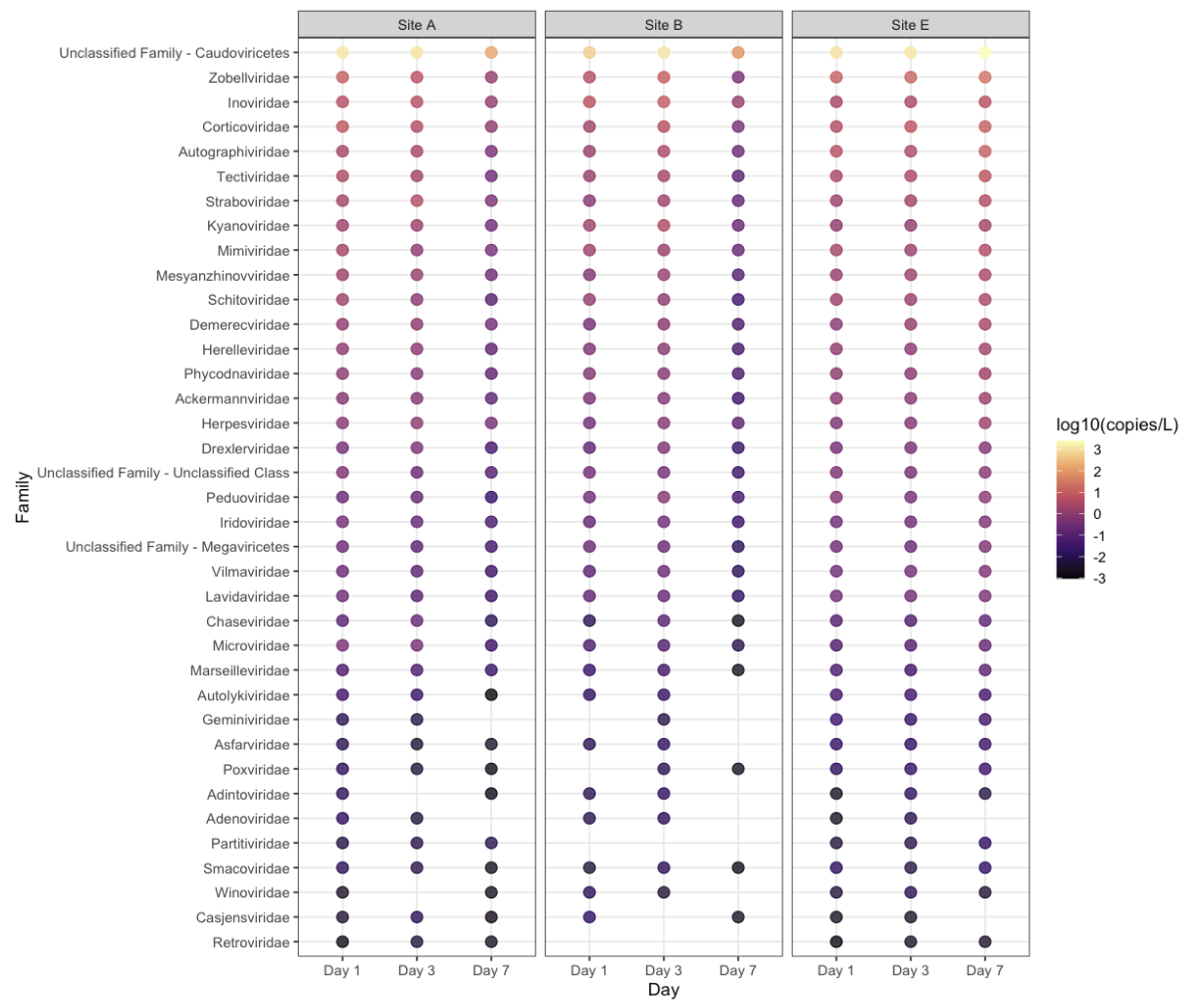

**Figure S4.** Virus families assigned to vOTUs detected in nine drinking water samples, with points colored based on the total vOTU concentration (log_10_-transformed copies/L) for each viral family. Families are ordered according to their total abundance across the samples. vOTUs unclassified at the family level were grouped by taxonomic class.

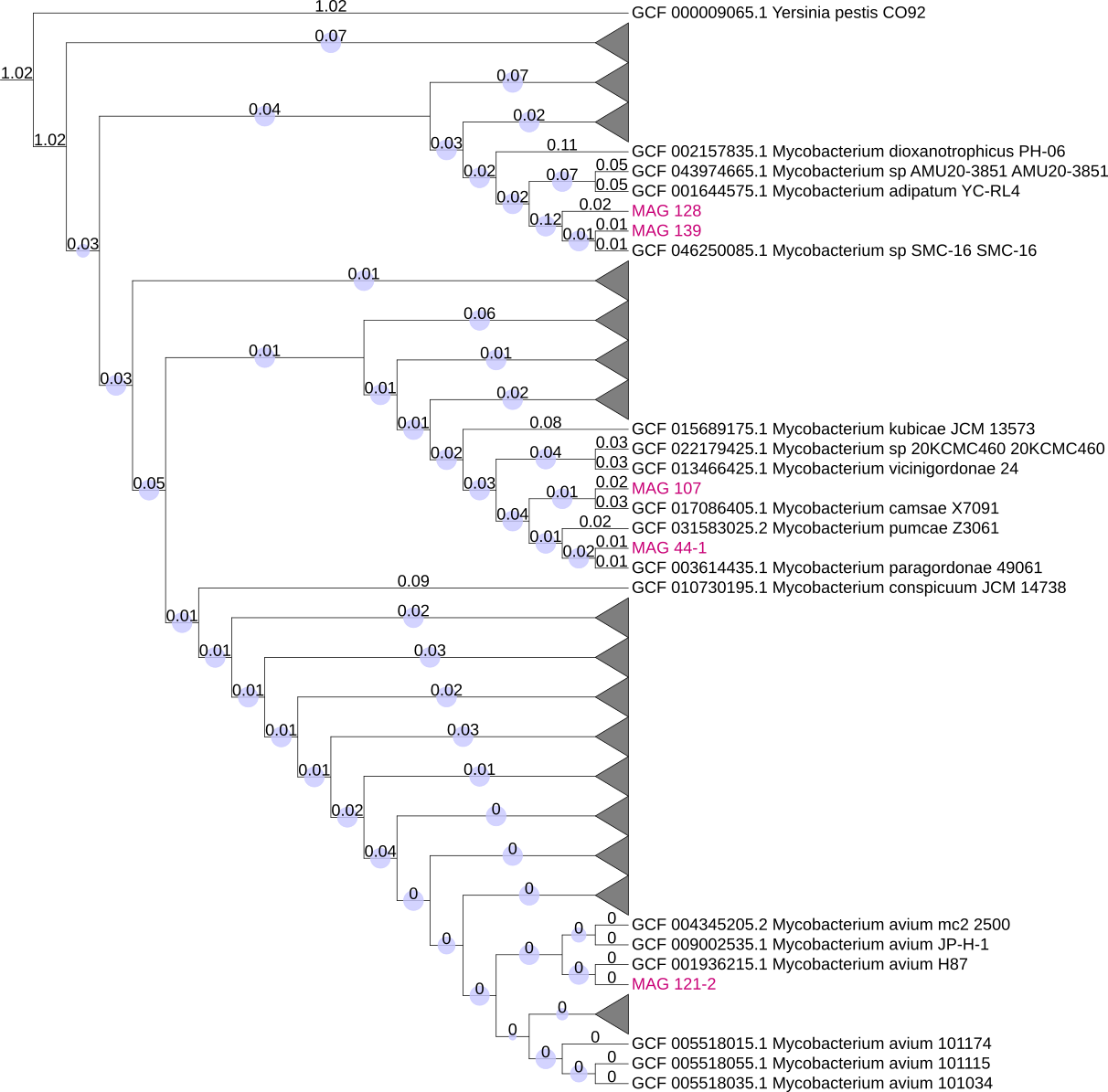

**Figure S5.** A maximum-likelihood phylogenetic tree was constructed using GToTree v1.8.6 (Lee, 2019) based on concatenated single-copy core genes identified across *Mycobacterium* reference genomes and NTM MAGs. The tree was rooted using *Yersinia pestis* as an outgroup. Reference genomes are labeled by NCBI accession numbers and species names, and MAGs are indicated by their assigned identifiers. Two of the seven NTM MAGs recovered in this study were excluded because they lacked sufficient single-copy marker genes required by GToTree. Black triangles represent collapsed reference NTM genomes that did not cluster with the MAGs recovered in this study.

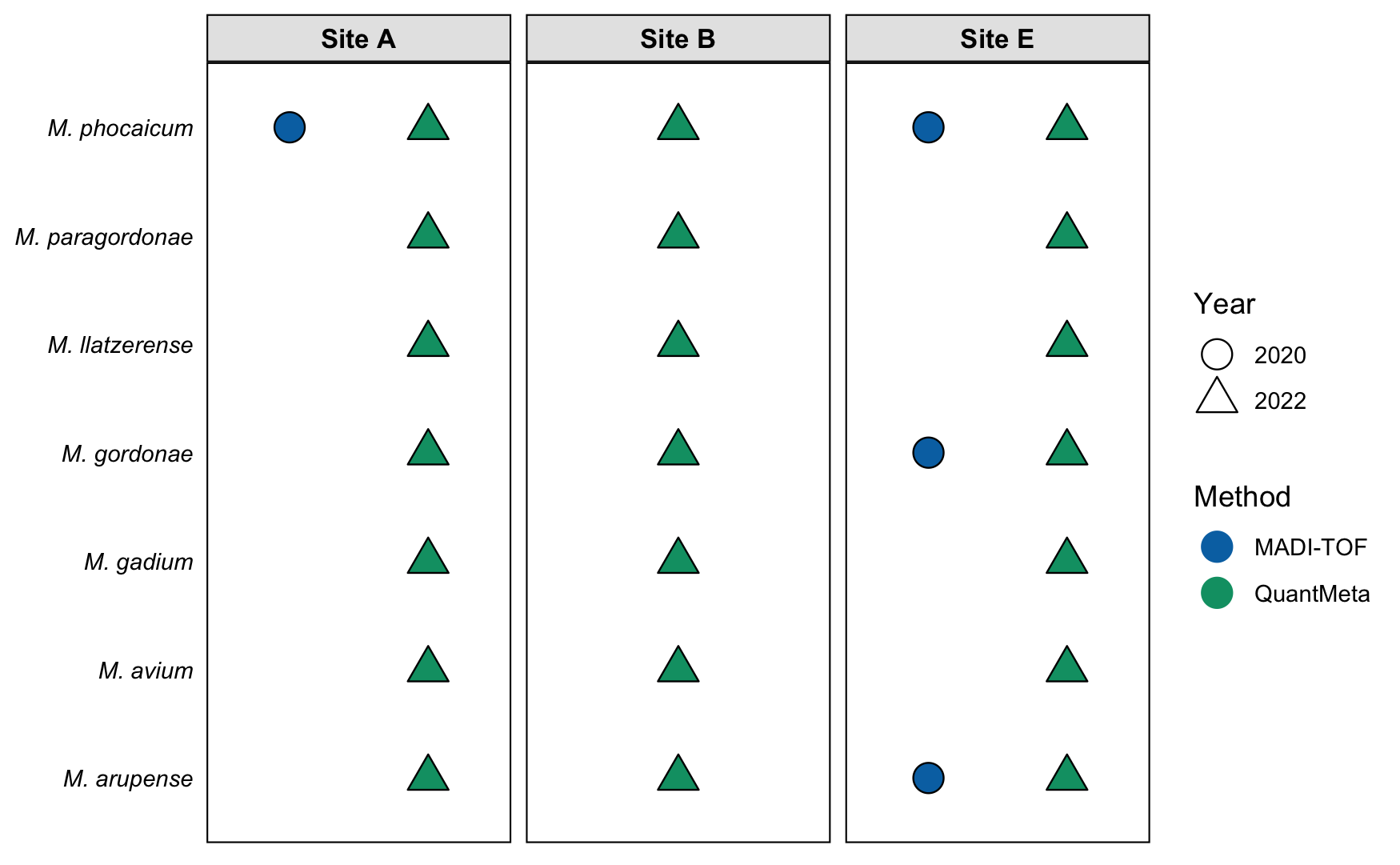

**Figure S6**. Detection of NTM species across three sample sites in two studies: Dowdell et al. (2024) analyzed NTM isolated from drinking water samples collected in 2020 using Matrix-Assisted Laser Desorption/Ionization–Time of Flight (MALDI-TOF) mass spectrometry. The current study collected samples in 2022 from the same sites and employed quantitative metagenomics (“QuantMeta”).

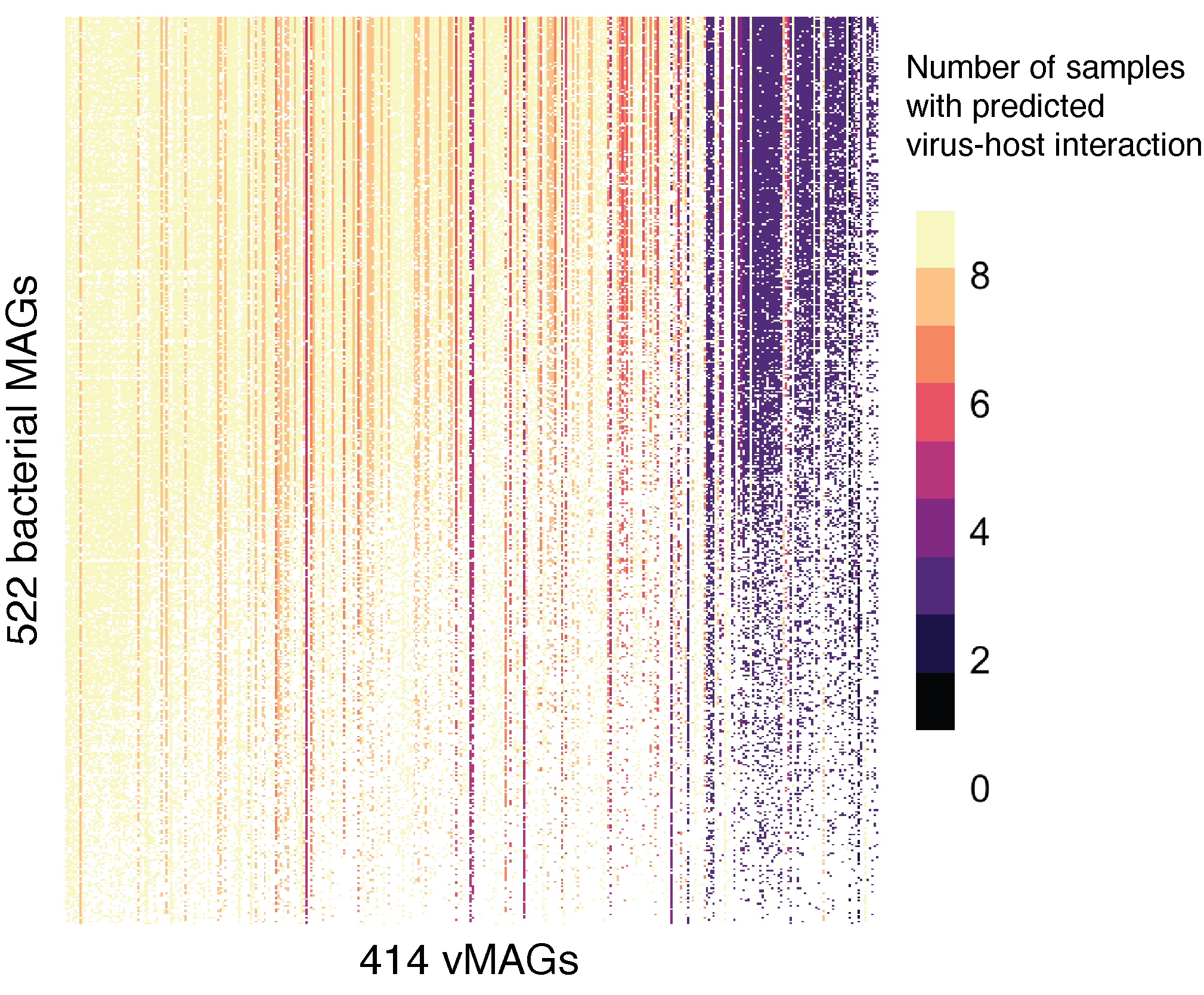

**Figure S7.** Overview of predicted viral–host interactions (VHIP) in nine drinking water samples, showing the network structure linking the 414 vMAGs (columns) to 522 bacterial MAGs (rows). Each connection represents a possible virus–host interaction as predicted by VHIP (Bastien et al., 2024), which is colored based on the number of samples in which the virus–host interaction was found (e.g., light yellow (9) represents pairs found in all nine samples, while black (0) represents pairs where infection was never predicted).

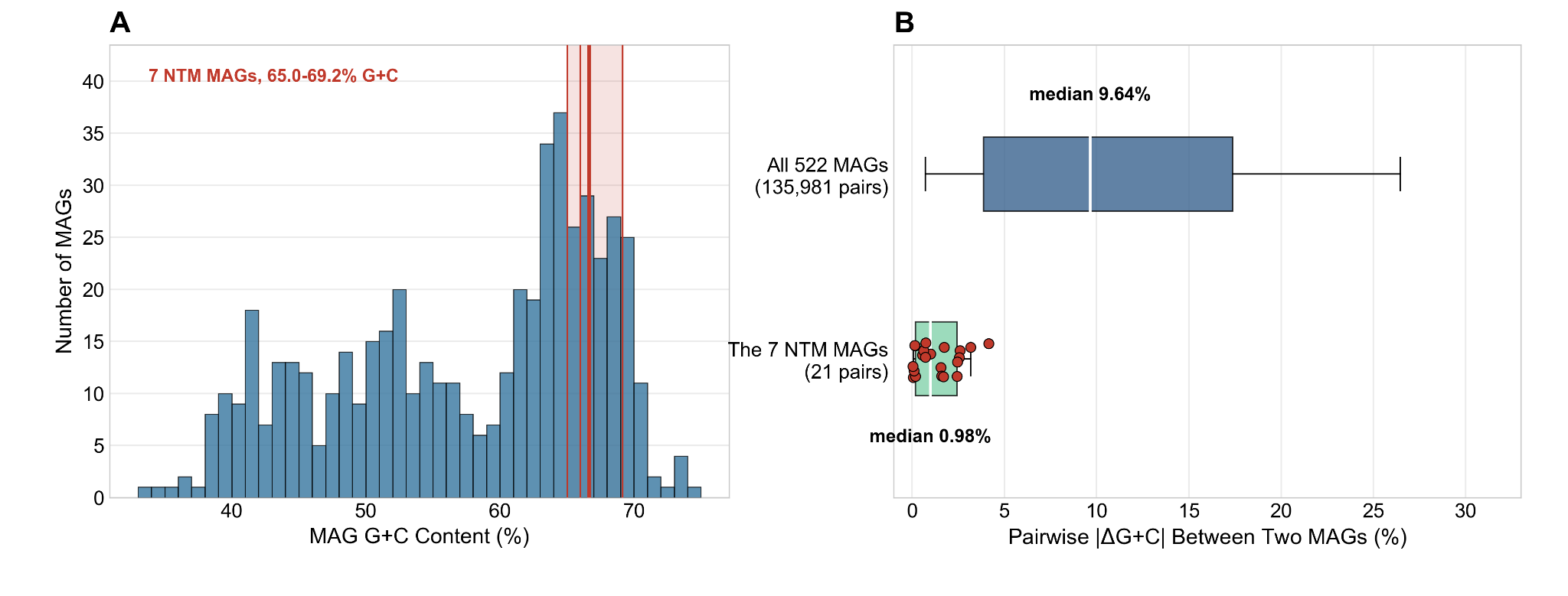

**Figure S8.** Genome GC composition of the bacterial MAGs. (A) GC content of the 522 bacterial MAGs, in 1% bins. Red lines mark the seven NTM MAGs, and the shaded band spans their range, 65.0–69.2%. (B) Pairwise absolute GC difference between two MAGs, for all 522 MAGs (135,981 pairs) and within the seven NTM MAGs (21 pairs, shown as points). Boxes span the interquartile range, whiskers the 5th and 95th percentiles, and the white line and the number beside each box are the median.

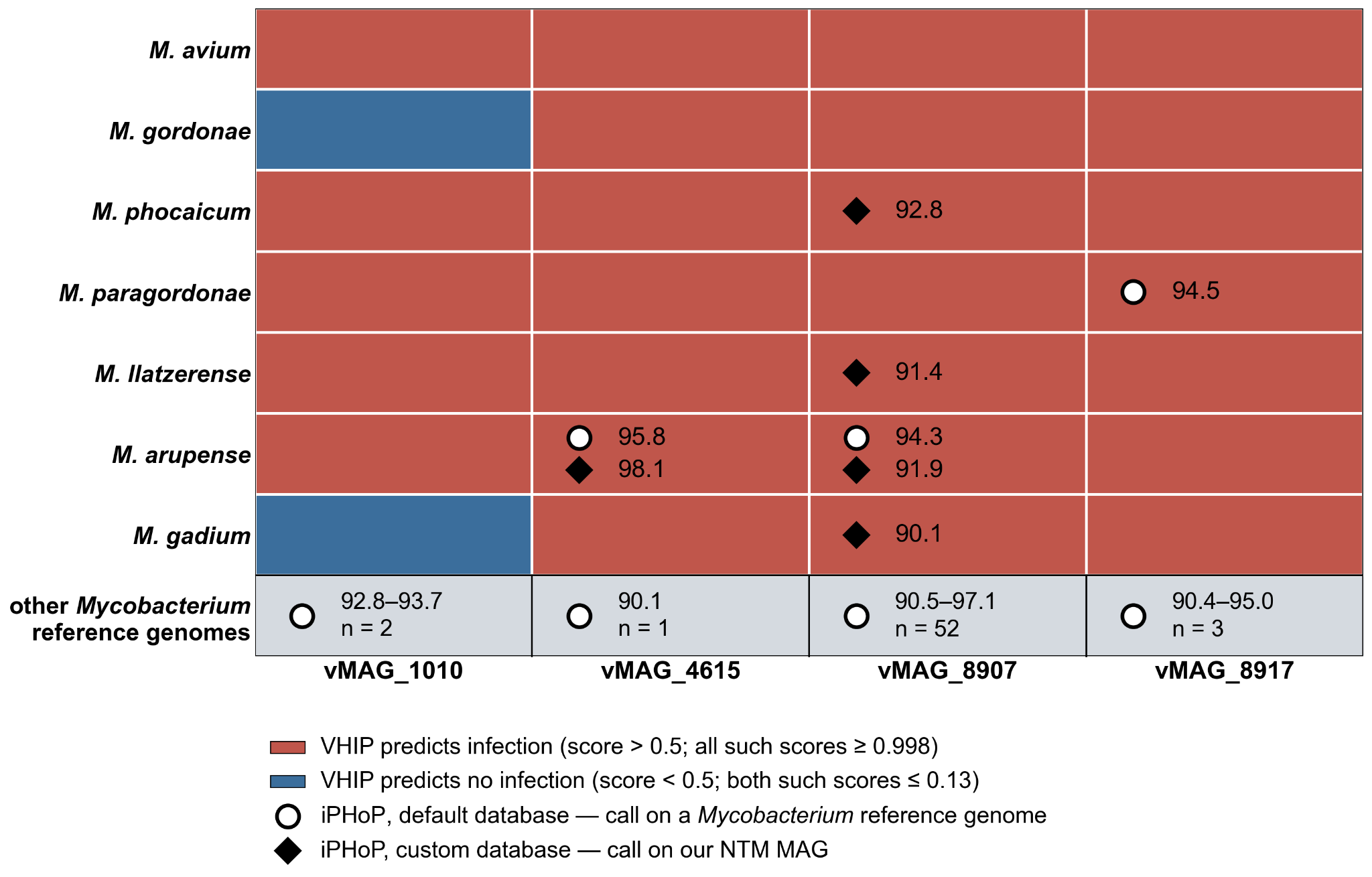

**Figure S9**. VHIP and iPHoP host predictions compared for the four vMAGs that iPHoP assigned to Mycobacterium. Columns are the four vMAGs and rows are the seven NTM MAGs, ordered by the number of vMAGs VHIP predicts for that MAG at a score of 0.99. Cell shading is the VHIP score for that vMAG–MAG pair, on a diverging scale with white at 0.5. Symbols inside a cell mark an iPHoP call, and the number beside each symbol is its confidence score. An open circle represents a call made with the default database, and a filled diamond represents a call made with the custom database containing our seven NTM MAGs. The bottom row gives the Mycobacterium reference genomes outside the seven species recovered here that iPHoP predicted to infect that vMAG. The iPHoP score range is listed with the number of predicted NTM host genomes below it.

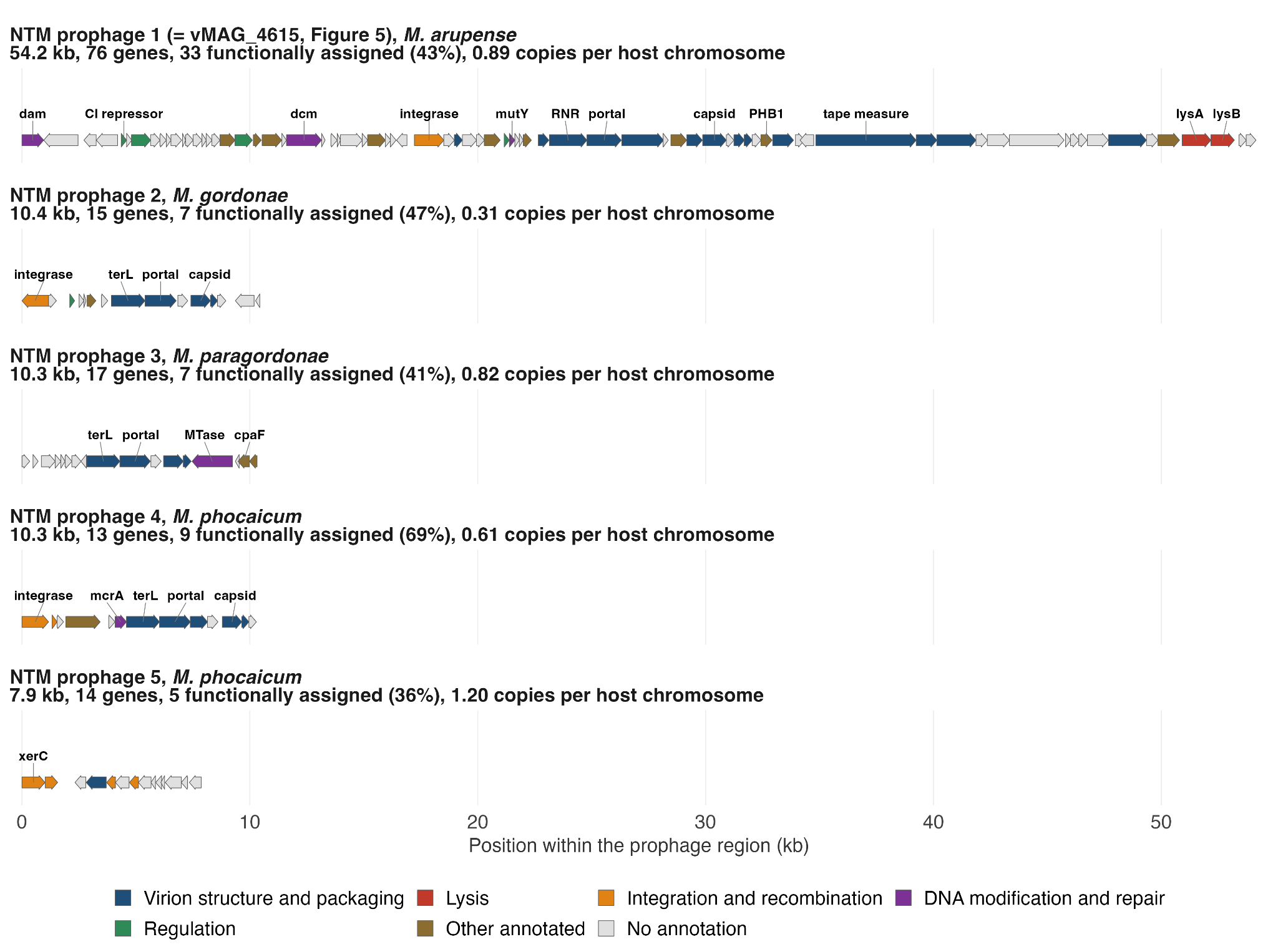

**Figure S10.** Gene maps of the prophage regions recovered from the NTM MAGs. Each arrow is a predicted gene and is colored by the functional category. Module assignments were made from combined annotations of geNomad, VIBRANT (Pfam and VOG), and GhostKOALA (KEGG Orthology), and genes matching only hypothetical proteins are shown as unannotated. Per-gene annotations are in Table S20. Strip headings provide the host MAG, the region length, the number of predicted genes, and the median copy number of the region per host chromosome across the nine samples.

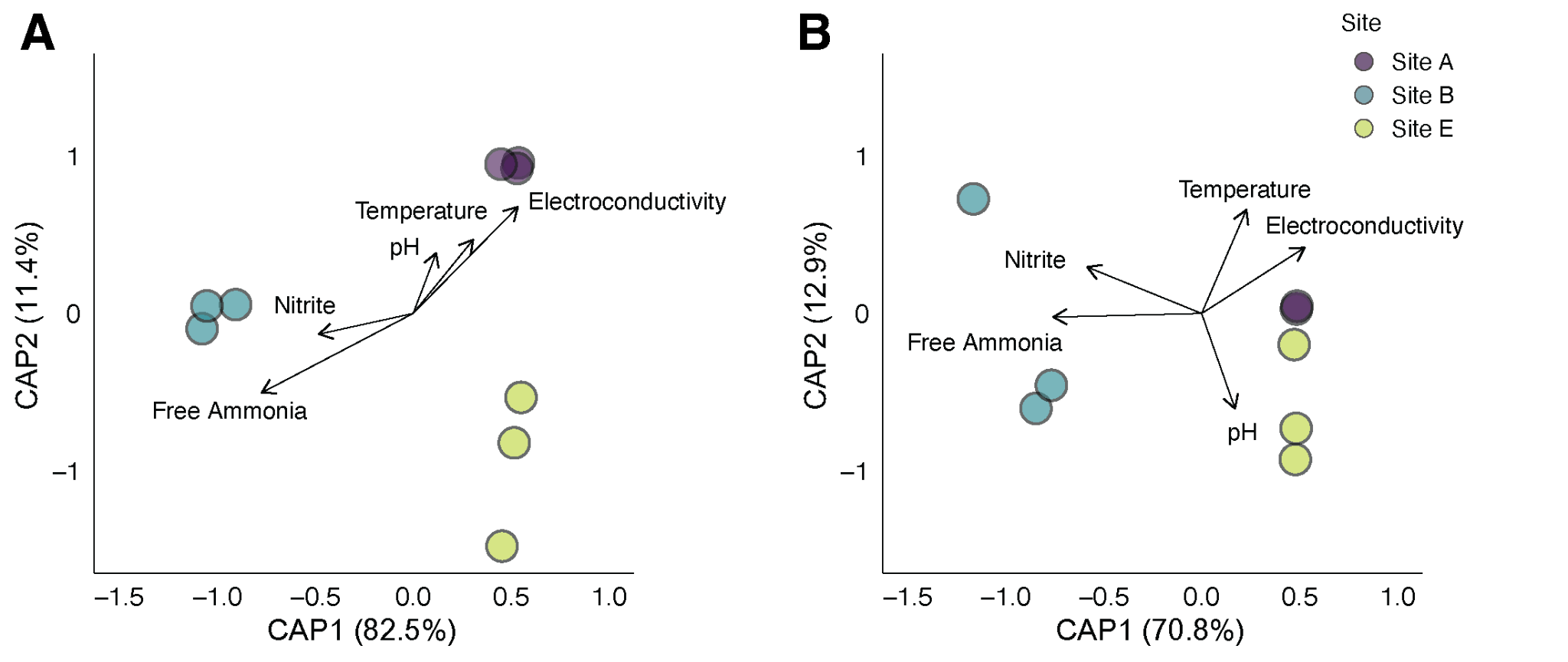

**Figure S11.** dbRDA plots showing (A) bacterial and (B) viral community structures in nine drinking water samples colored by sample site. Ordinations were generated using Hellinger-transformed read counts. Vectors indicate water chemistry characteristics significantly associated with community variation.

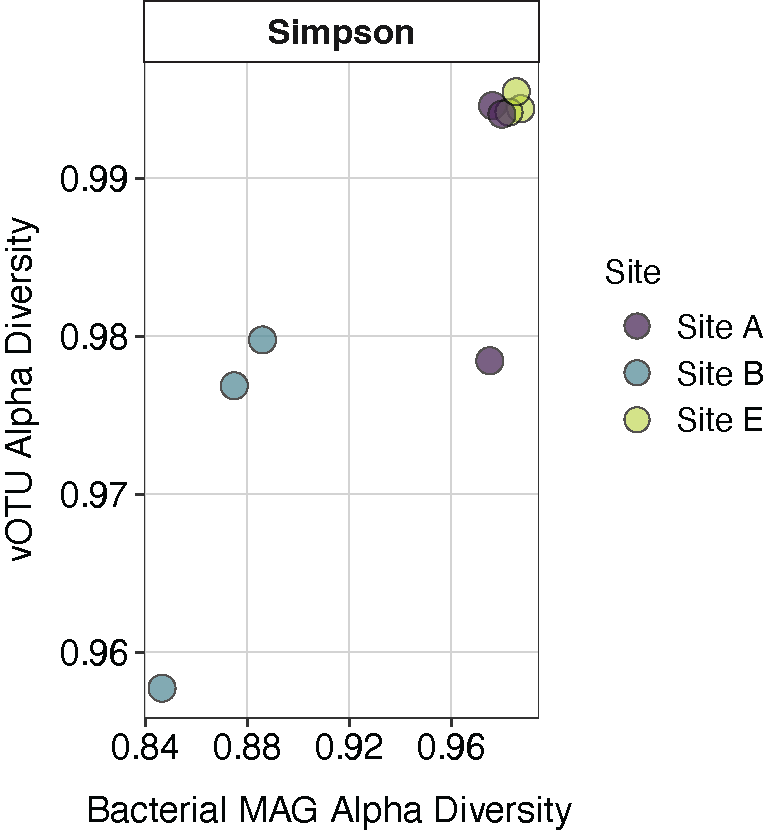

**Figure S12**. Simpson index of bacterial and viral communities in nine drinking water samples, colored by sample site.

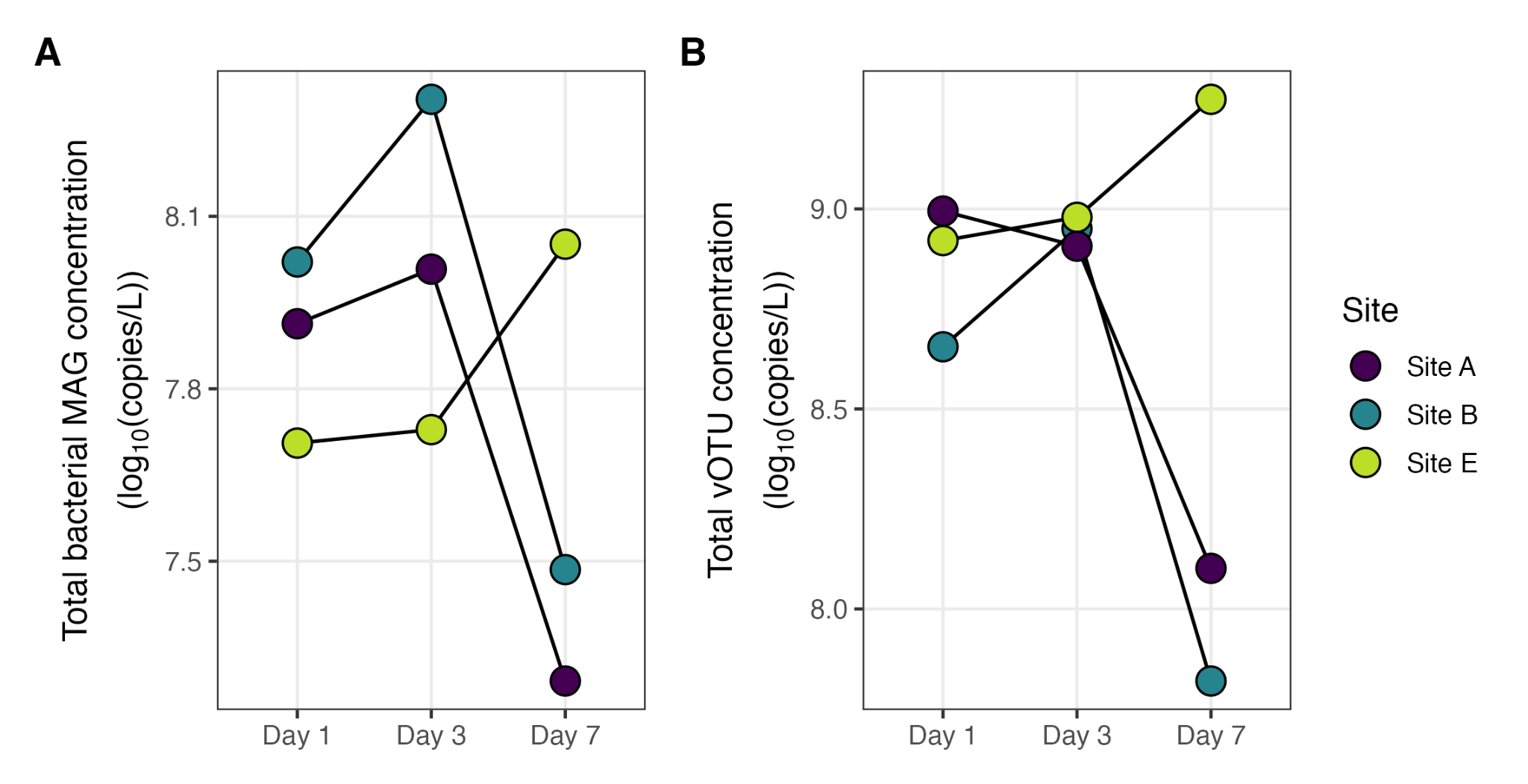

**Figure S13**. Total bacterial MAG and vOTU concentration in nine drinking water samples, colored by sample site.

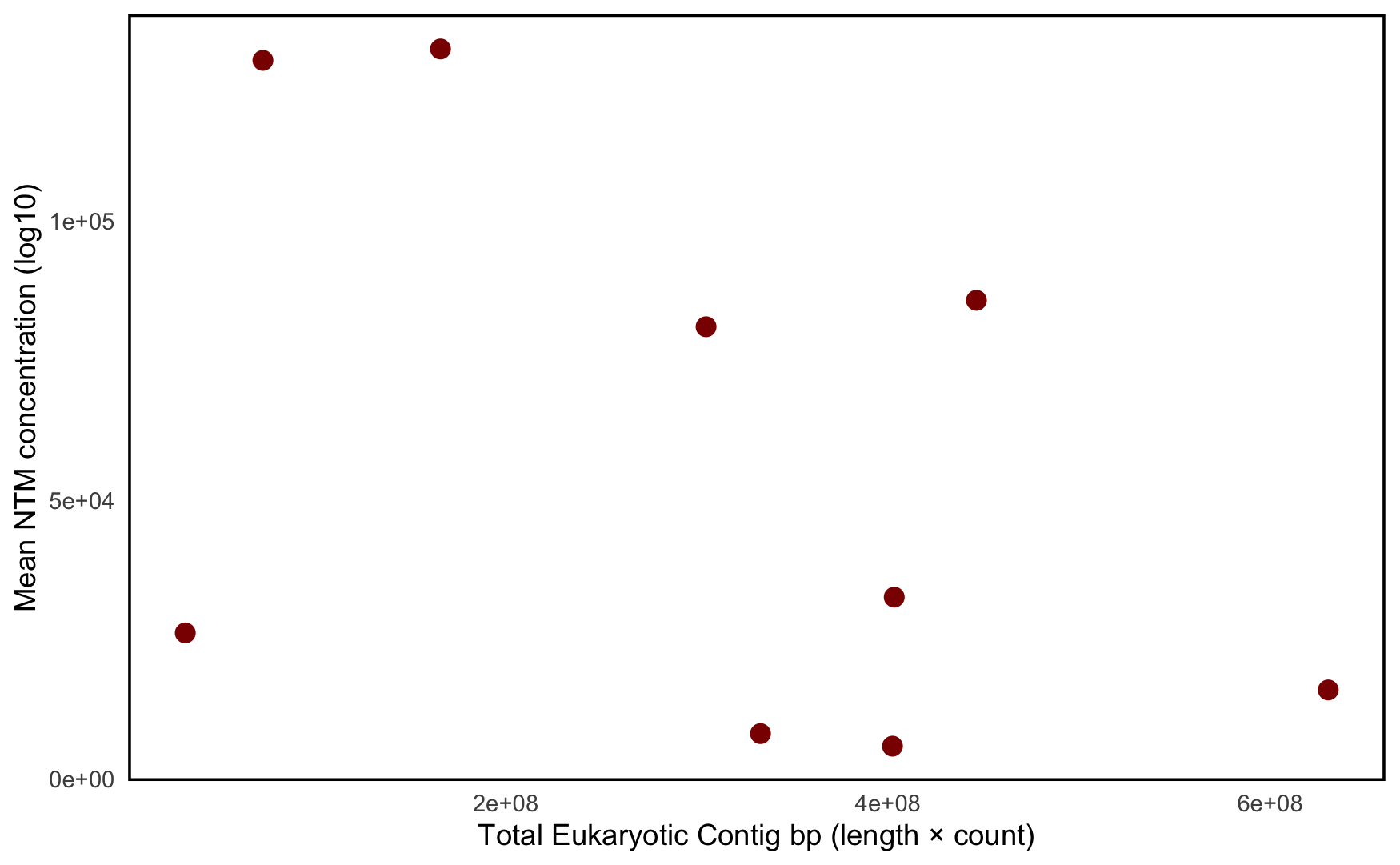

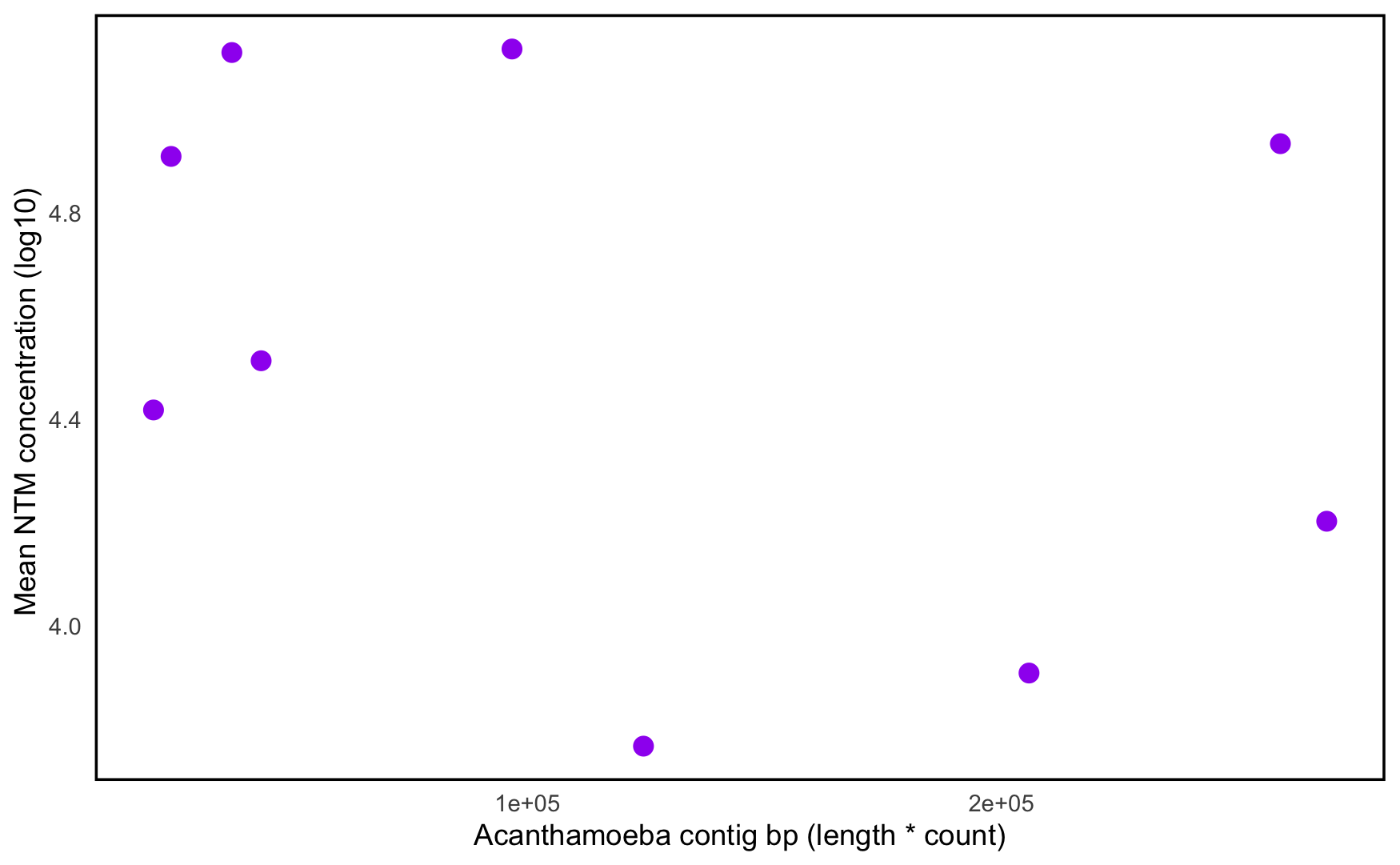

**Figure S14.** Relationship between (A) eukaryotic contig base pairs (contig length × count) and the average concentration of NTM MAGs (log_10_-transformed copies/L), and (B) *Acanthamoeba* contig base pairs and the average concentration of NTM MAGs (log_10_-transformed copies/L).

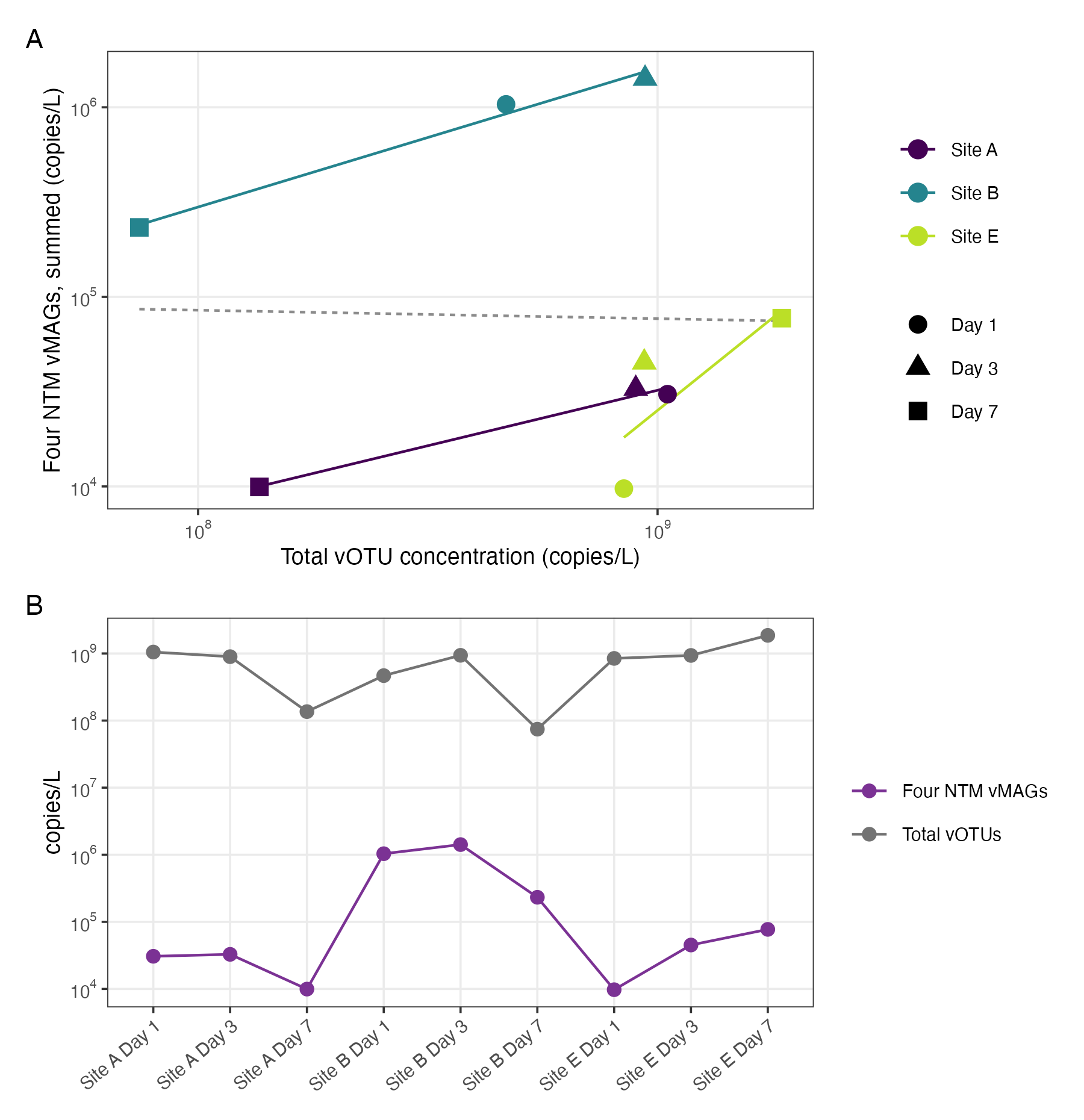

**Figure S15.** Total concentration of the four vMAGs (vMAG_1010, vMAG_4615, vMAG_8907, and vMAG_8917) vs. the total vOTU concentration across the nine first-draw drinking water samples.

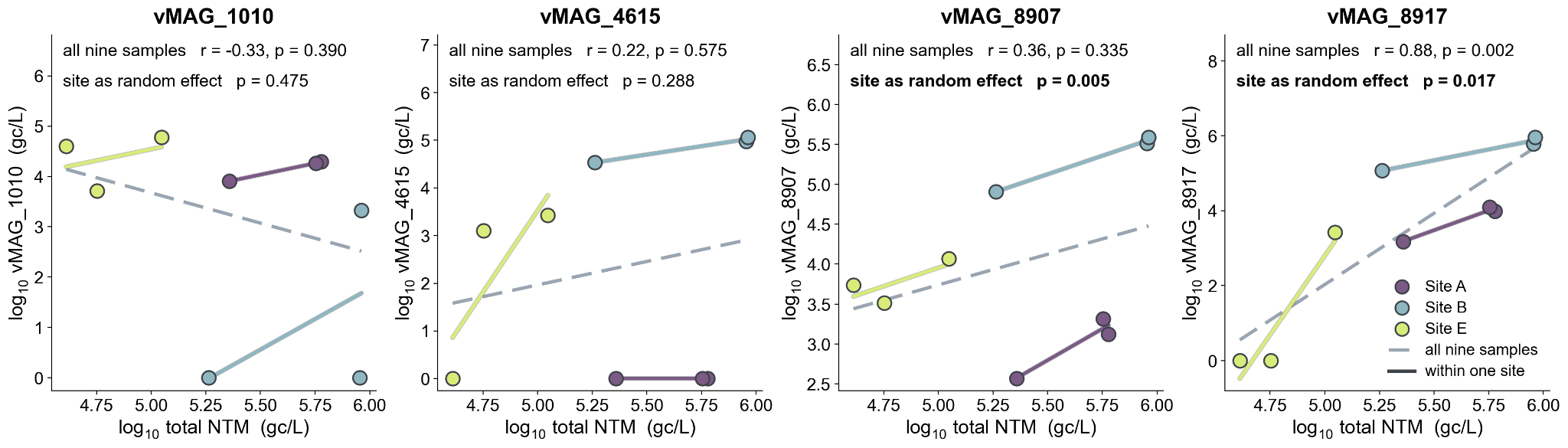

**Figure S16.** Concentration of each of the four vMAGs against total NTM concentration in the same sample, within and across sites. Each panel is one vMAG across the nine samples, colored by site; the y-axis differs between panels. The dashed grey line is the regression across all nine samples, and the solid colored lines are separate regressions within each site (three samples per site). Each panel reports the Pearson correlation across all nine samples and, below it, the p-value for the slope when site is fitted as a random effect (linear mixed-effects model of log_10_ vMAG concentration on log10 total NTM concentration), in bold where p < 0.05. vMAG concentrations are log_10_(copies/L + 1), so a value of zero means the vMAG was not detected in that sample.

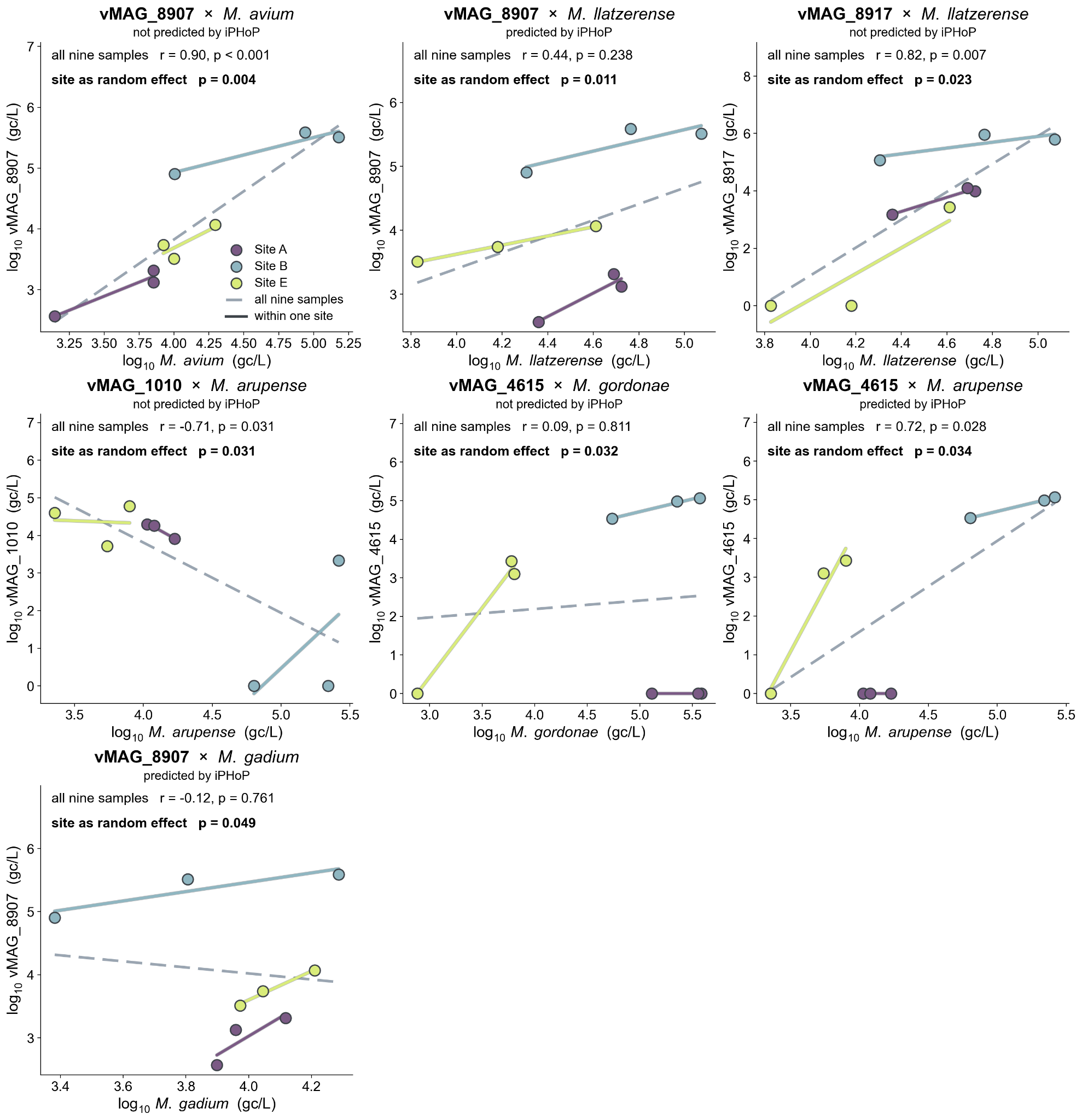

**Figure S17.** Concentration of individual vMAGs against individual NTM MAGs, for the pairs with a significant site-adjusted association. A nominally significant correlation in abundance was found for seven out of the 26 vMAG–NTM pairs that VHIP predicted at a score of 0.99 among the four candidate NTM-associated vMAGs and the seven NTM MAGs (*p* < 0.05 in a linear mixed model fitted with site as a random intercept). Panels are ordered by increasing p-value, and the selection is post-hoc, so the remaining 19 pairs are not shown; none of the 26 pairs remained significant after Benjamini–Hochberg correction across all 26 (minimum q = 0.11). Each panel shows the nine samples, colored by site, with gene concentrations log_10_-transformed on both axes. The dashed grey line is the regression over all nine samples, and the solid lines are regressions within each site. The first line in each panel shows the Pearson correlation and its p-value across all nine samples, and the bold line shows the p-value from the mixed model.

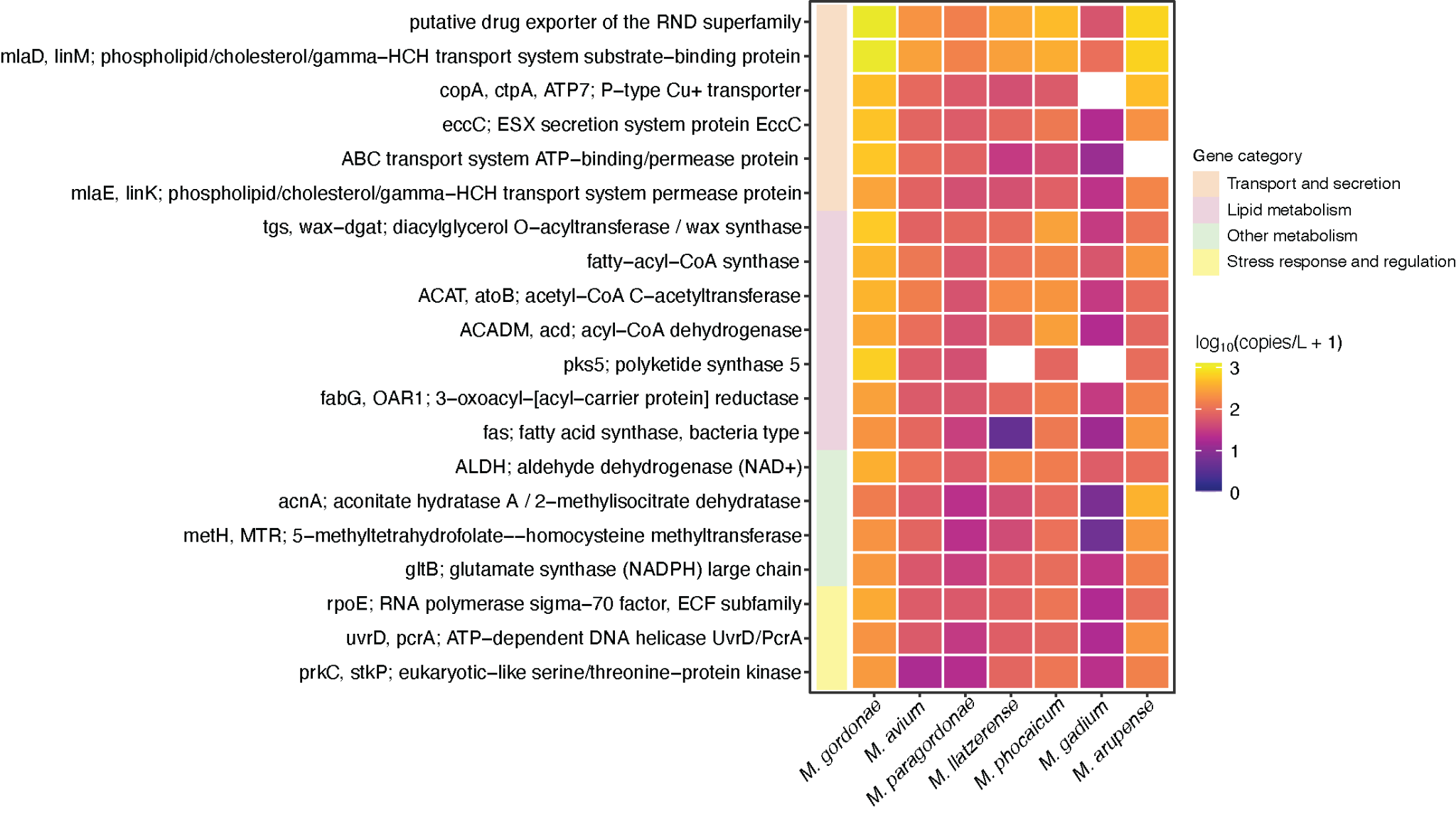

**Figure S18.** Metabolic potential of NTM MAGs. The 20 KEGG Orthology IDs with the highest overall mean absolute concentrations across the seven NTM MAGs are shown. For each MAG, gene concentrations assigned to the same KO were summed within each sample and averaged across the nine samples; KOs absent from a MAG were assigned a value of zero. Heatmap color represents log₁₀-transformed mean KO concentration for each MAG (log_10_ copies/L+1). The color coding in the first column indicates the KO functional category.

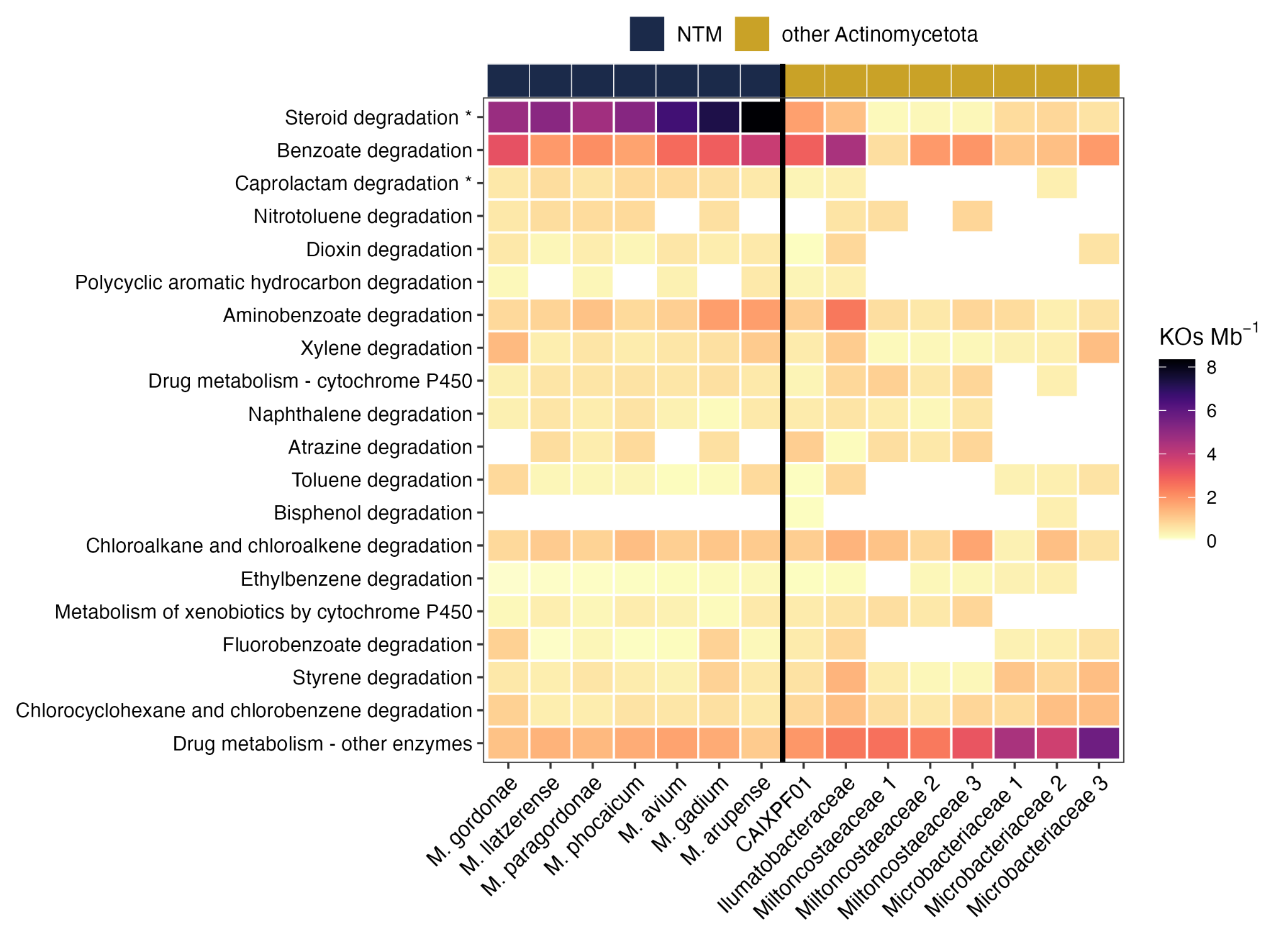

**Figure S19.** Xenobiotic biodegradation pathways across NTM and co-occurring Actinomycetota MAGs. The 20 KEGG level-C pathways within the xenobiotic biodegradation and metabolism category are shown, expressed as KEGG Orthology IDs (KOs) per Mb of assembly. Pathways are ordered by the difference between group medians. Asterisks indicate the two pathways significantly enriched in NTM relative to the other Actinomycetota after Benjamini–Hochberg correction (q < 0.05).

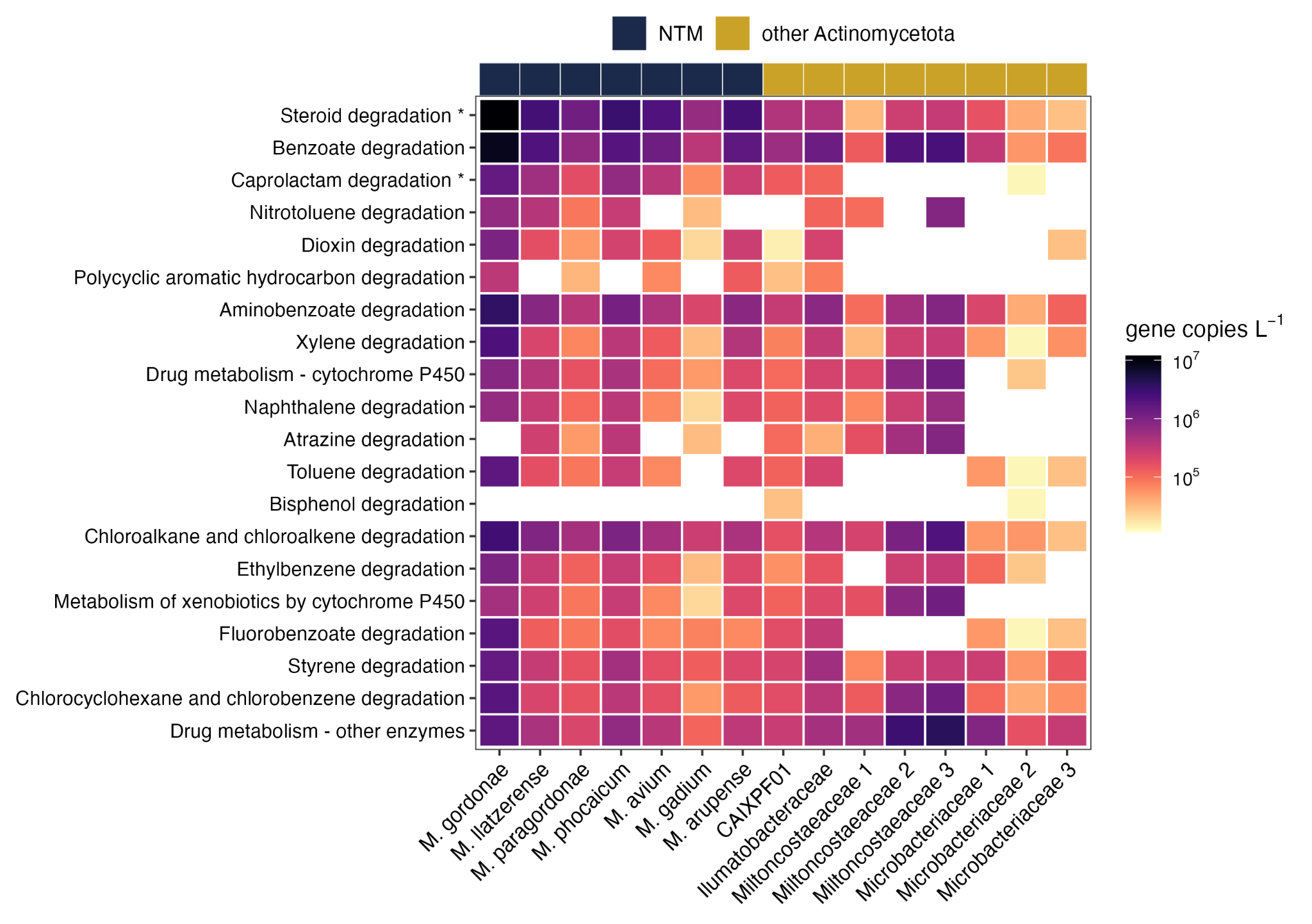

**Figure S20.** Xenobiotic biodegradation and metabolism gene concentrations across the seven NTM MAGs and the eight other Actinomycetota MAGs recovered in this study. Rows are the 20 KEGG level-C pathways within the category; columns are individual MAGs, colored along the top by group (NTM or other Actinomycetota MAGs). Cell color represents gene concentration on a log_10_ scale, averaged across the nine samples; white indicates that no genes in that pathway were detected in that MAG. Asterisks mark the pathways for which the NTM MAGs were higher than the other Actinomycetota (Wilcoxon rank-sum with Benjamini–Hochberg correction across the 20 pathways, q < 0.05).

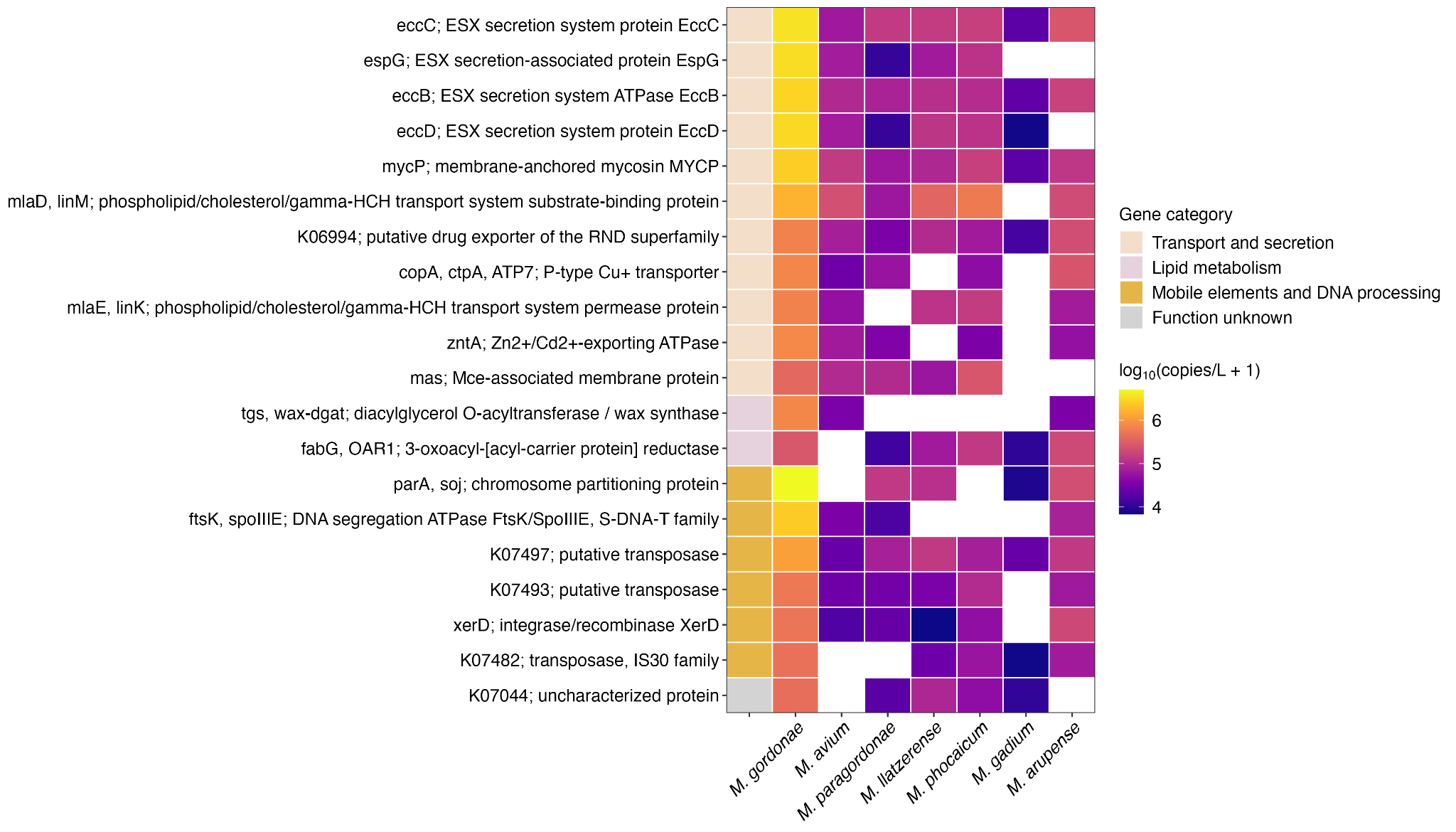

**Figure S21.** The 20 KEGG Orthology IDs (KOs) with the highest overall mean absolute concentrations were identified in plasmid-associated contigs from the seven NTM MAGs. Gene concentrations assigned to the same KO were summed within each sample and MAG, then averaged across the nine samples. KO–MAG combinations absent from a given MAG were assigned a value of zero. Heatmap colors represent the log_10_-transformed mean KO concentration for each MAG [log_10_(copies/L + 1)], and the first column indicates the functional category assigned to each KO.

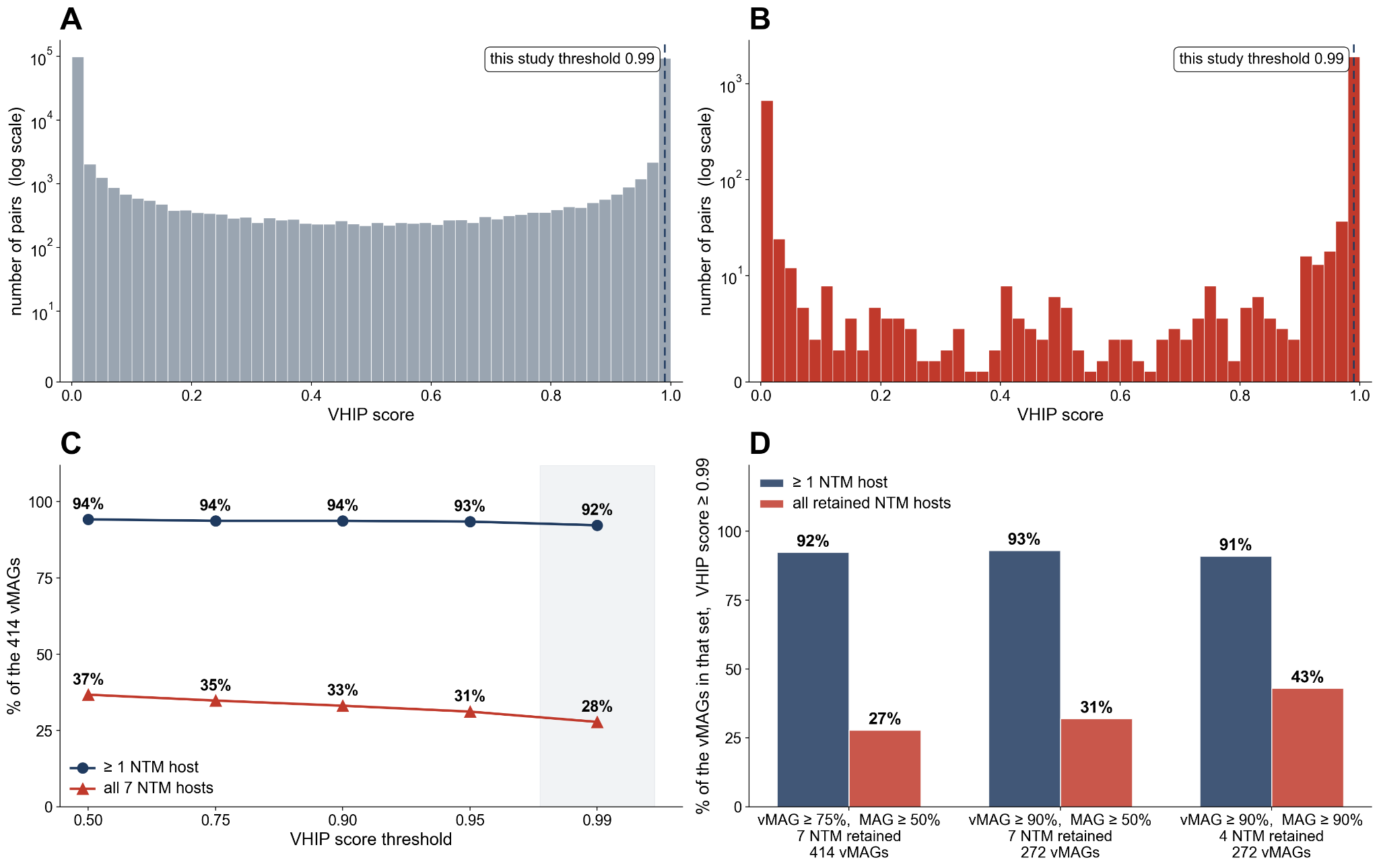

**Figure S22.** (A) Distribution of VHIP scores for all 216,108 vMAG–bacterial MAG pairs (414 vMAGs against 522 MAGs) and (B) for the 2,898 vMAG–NTM MAG pairs, on a log-scaled y-axis. Each pair appears once, at its highest score across the per-sample networks. The dashed line marks the score of 0.99 used in this study. (C) Percentage of the 414 vMAGs predicted to interact with at least one of the seven NTM MAGs, and with all seven, at score thresholds from 0.50 to 0.99. The shaded band marks the 0.99 threshold reported throughout. (D) The same two percentages at a score threshold of 0.99, for three combinations of viral and bacterial genome completeness: the thresholds used here (vMAG ≥ 75%, MAG ≥ 50%; 414 vMAGs against 522 MAGs), a stricter viral threshold (vMAG ≥ 90%, MAG ≥ 50%; 272 vMAGs against 522 MAGs), and both thresholds raised (vMAG ≥ 90%, MAG ≥ 90%; 272 vMAGs against 299 MAGs).

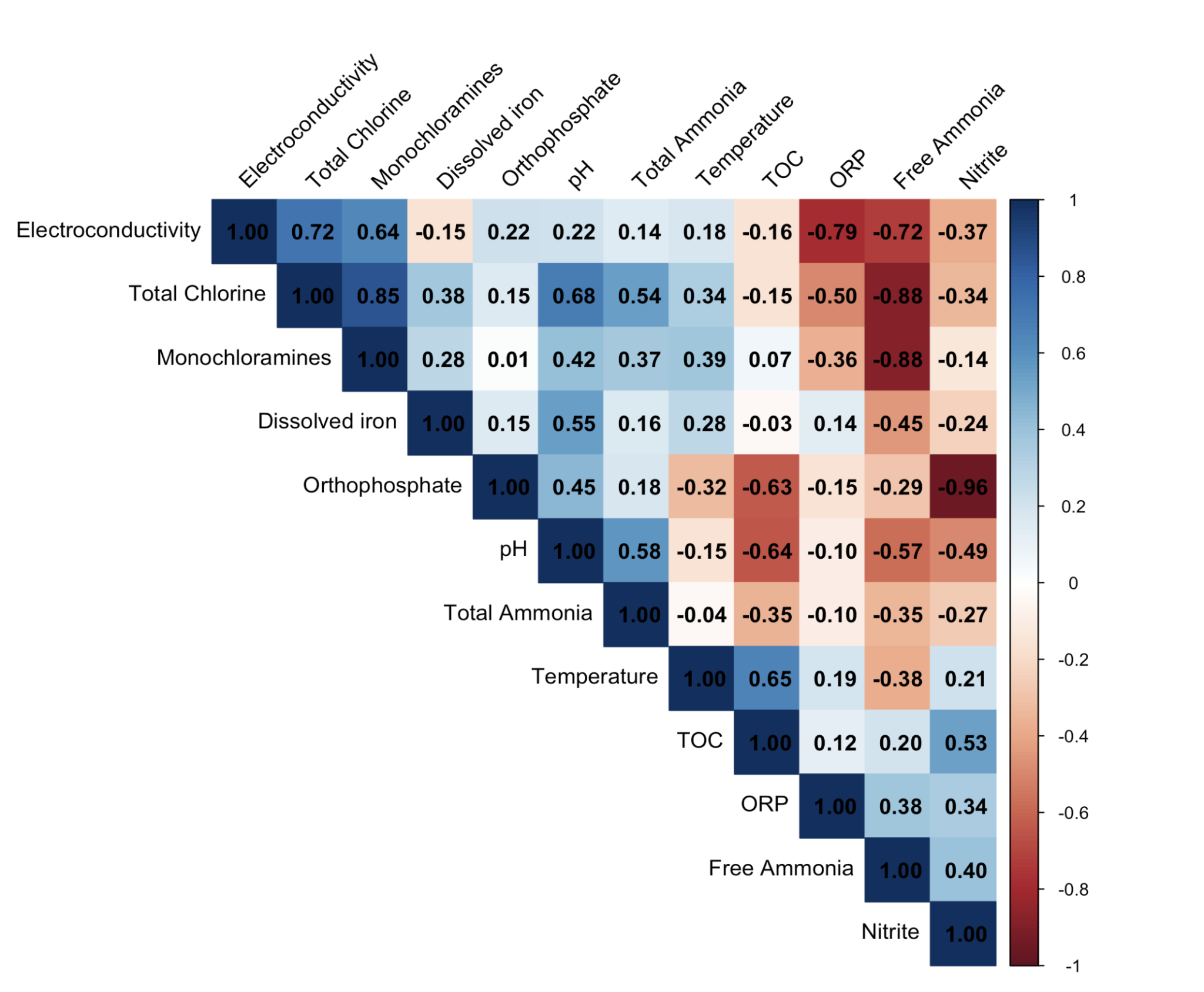

**Figure S23.** Pearson correlation matrix showing pairwise correlations among physicochemical parameters measured in nine drinking water samples.
